# Accounting for gene action improves the accuracy of polygenic scores for some complex traits

**DOI:** 10.64898/2026.10.15.26365665

**Authors:** Yuxuan Sun, Fabio Morgante, Trudy F.C. Mackay

## Abstract

Polygenic scores (PGS) predict an individual’s genetic value for a complex trait of interest by aggregating effect size estimates from genome-wide association studies (GWAS). While powerful, this approach assumes that genetic variants affect the phenotype in an additive way, which might be problematic when dominance is part of the genetic architecture. To address this limitation, we developed PGS_*a,d*_, a simple polygenic scoring framework, rooted in quantitative genetics theory, that incorporates both additive (*a*) and dominance effects (*d*) directly. We evaluated PGS_*a,d*_ against the standard GWAS-based PGS (PGS_*β*_) in simulations. The results showed that PGS_*a,d*_ consistently achieved lower root mean squared error (RMSE) and progressively higher *R*^2^ as dominance contributed more to the genetic architecture. We then applied the method to 16 blood cell traits, height, and body mass index (BMI) from UK Biobank. PGS_*a,d*_ showed comparable *R*^2^ to PGS_*β*_ for all traits (including height and BMI whose architecture is predominantly additive) and significantly lower RMSE for four traits after strict Bonferroni correction. Across traits, RMSE improvement from PGS_*a,d*_ was positively associated with the magnitude of the estimated dominance effects, whereby traits with larger dominance effects displayed lower RMSE from PGS_*a,d*_. As a byproduct of our analysis, we also identified 141 variants in 39 genomic regions with significant dominance effects after Bonferroni correction across traits, recapitulating known associations and identifying novel ones. Together, these results show that explicitly modeling additive and dominance effects is a robust strategy that can improve individual-level predictions, especially when dominance contributes to complex trait architecture.

## Introduction

Complex traits are affected by genetic and non-genetic factors, and encompass medically-relevant and agriculturally-important traits [1, 2]. Thus, understanding the genetic architecture of complex traits has important implications for precision medicine, agriculture, and evolutionary biology. Quantitative genetics theory has given us the means to achieve this goal.

### Quantitative genetics theory

The first attempt at deriving a mathematical model describing how genetic and non-genetic (or environmental) factors determine a complex phenotype was made by Fisher [3], who introduced the standard quantitative genetic model:

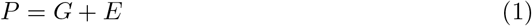

where *P* is the phenotypic value, *G* is the genotypic value (which includes the effects of all the variants affecting the trait), and *E* is the environmental deviation (which includes the effects of all the environmental factors on the trait) [4].

The effect of a single locus (A) with two alleles (A_1_ and A_2_) affecting the trait of interest is modeled as follows [4]. Assuming that A_1_ is the allele increasing the trait value and the population’s mean environmental deviation is zero, we convert the means of the phenotypic values for each of the three genotypes to genotypic effects on an arbitrary scale depicted in Figure 1, where the midpoint between the phenotypic means of the two homozygotes defines the origin (0). On this scale, the genotypic effects of A_1_A_1_ and A_2_A_2_ are denoted as +*a* and −*a*, respectively, where *a* represents the *additive effect* of the variant and can be calculated as:

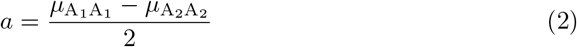

where *µ*_*xx*_ represents the mean phenotype of the *xx* genotype. The genotypic effect of *A*_1_*A*_2_ is expressed as a deviation from the origin, is denoted as *d*, and can be calculated as:

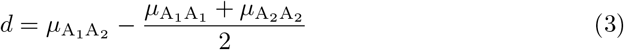

**Fig 1.**
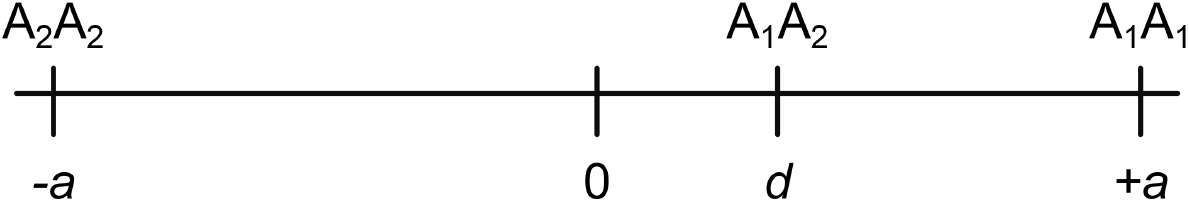
Illustration of additive and dominance effects on an arbitrary scale.

Under this parameterization, *d* = 0 corresponds to purely additive gene action, −*a < d <* 0 indicates partial dominance of the A_2_ allele, 0 *< d <* +*a* indicates partial dominance of the A_1_ allele, *d* = −*a* indicates complete dominance of the A_2_ allele, *d* = +*a* indicates complete dominance of the A_1_ allele, *d <* −*a* indicates under-dominance, and *d >* +*a* indicates over-dominance. Thus, *d* is the *dominance effect* of the variant.

This formulation makes it clear that *a and d are properties of genotypes*. However, genotypes are segregated at meiosis and alleles are transmitted to the next generation according to Mendelian expectation. Thus, assuming an ideal population satisfying some assumptions (*e*.*g*., random mating, no evolutionary forces), Fisher derived the *average effects (α*_1_ *and α*_2_*) as population measures of the effects of individual alleles* on a phenotype, which are defined as the mean value of all progeny that received that allele from a parent, the other allele having come at random from the population, expressed as a deviation from the overall population mean.

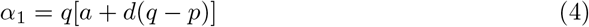

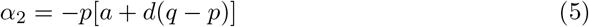

where *p* and *q* are the frequencies of the A_1_ and A_2_ alleles (*p* + *q* = 1), respectively. From the average effects, we can compute the average substitution effect, which is denoted as *α* and is defined as the effect of randomly replacing an A_2_ allele with an A_1_ allele:

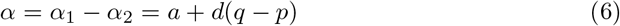

Having defined the effects of individual alleles, we can now express the value of a genotype as the sum of the average effects of the allele it carries. This is called the breeding value or additive genetic value (*A*) of a genotype and can be calculated as follows:

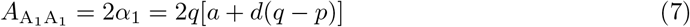

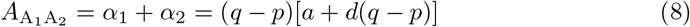

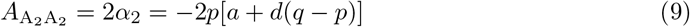

Importantly, the additive genetic value of a genotype does not represent the full genotypic value (*G*), defined as the genotypic effect minus the population mean:

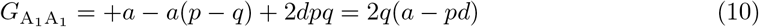

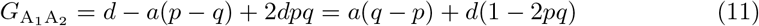

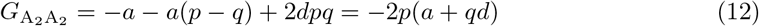

In fact, the additive genetic value of the heterozygote is constrained to be halfway between the additive genetic values of the homozygotes and, hence, does not allow for dominance. The additive genetic value is of fundamental importance in quantitative genetics – especially in agricultural breeding – because it provides a measure of the part of the genotypic value that a parent can transmit to its progeny (alleles and not genotypes are transmitted to the next generation). The difference between the genotypic value and the additive genetic value is called the *dominance deviation* (*D*) and accounts for the potential non-linearity of the genotypic value induced by dominance.

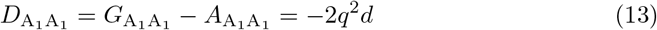

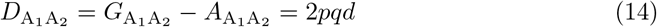

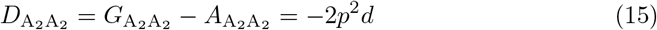

The relationship between all these parameters is shown in Figure 2, which depicts the regression of genotypic effects on genotypes – coded as number of copies of A_1_ – weighted by genotype frequencies. This figure highlights that the slope of this regression is the average substitution effect (*α*), and the linear (*i*.*e*., additive) relationship between the additive genetic values and genotypes, with the former lying on the regression line.

**Fig 2.**
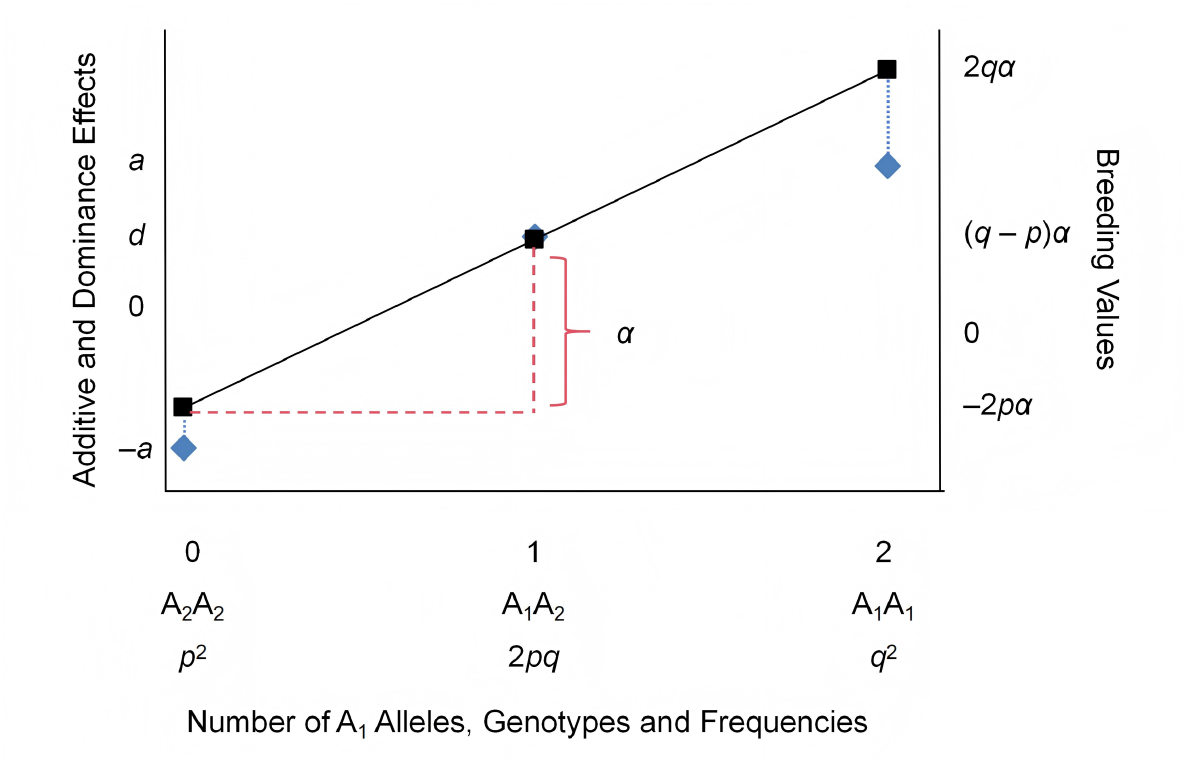
Relationships between average effects, genotype and breeding values. An illustrative example is shown for a locus with *a* = 1, *d* = 0.2, *p* = 0.4 for allele A_1_ and *q* = 0.6 for allele A_2_. The genotypic effects of A_2_A_2_, A_1_A_2_, and A_1_A_1_, denoted by −*a, d*, and *a*, respectively, are regressed on the number of copies of the A_1_ allele. The slope of this linear regression is the average effect of allele substitution, *α*, as illustrated by the red dashed lines. The blue diamonds represent the genotypic values of the three genotypes, whereas the black squares represent their corresponding breeding values. The departures of breeding values from genotypic value represented by the blue dashed line are the dominance deviations.

It is very important to realize the difference between *effects* (*a* and *d*) and *values* (*A* and *D*). Effects refer to gene action and do not depend on allele frequencies. On the other hand, values are properties of genotypes in a randomly mating population and are frequency-dependent. In addition, there is not a 1-to-1 relationship between effects and values. In fact, as we can see from the expressions above, while the additive effect contributes only to additive genetic values, the dominance effect contributes to both additive genetic values and dominance deviations.

One of the goals of quantitative genetics is to relate the observed phenotypic variation to components due to different sources. Thus, in its simplest form, the quantitative genetic model at the level of variation is:

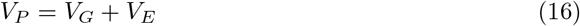

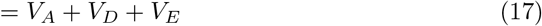

where *V*_*P*_ is the variance of the phenotypic values or phenotypic variance, *V*_*G*_ is the variance of the genotypic values or genotypic variance, *V*_*A*_ is the variance of the additive genetic values or additive variance, *V*_*D*_ is the variance of the dominance deviations or dominance variance, and *V*_*E*_ is the variance of the environmental deviations or environmental variance. The genetic variance components are computed as:

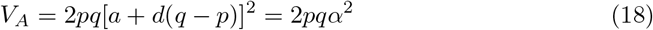

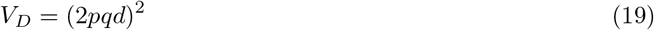

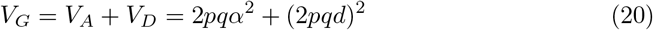

Again, these expressions show the lack of 1-to-1 relationship between effects and genetic variance components – note that the dominance effect contributes to both additive and dominance variance components.

Broad sense heritability (*H*^2^) and narrow sense heritability (*h*^2^) can be computed as ratios of variance components:

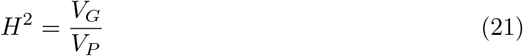

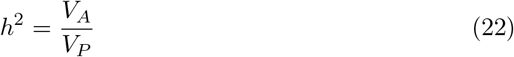

### Uncovering variants affecting complex traits and polygenic prediction

Initially, investigating the genetic architecture of complex traits mostly meant estimating variance components and heritability, which can be done without knowing the actual variants affecting the trait and their effects and frequencies [4, 5]. With the development of inexpensive genotyping arrays and the decreasing cost of genome sequencing, we have been able to genotype thousands of individuals at hundreds of thousands or even millions of genetic variants. This has given us the possibility to find genetic variants associated with complex traits with Genome-Wide Association Studies (GWAS) [6].

GWAS typically test the association between a variant and a phenotype using an additive allele-dosage regression model. Using the same notation as above, for a biallelic locus with alleles A_1_ and A_2_, let *x* denote the number of copies of a reference allele (A_1_) such that *x* = 0, *x* = 1, and *x* = 2 correspond to A_2_A_2_, A_1_A_2_, and A_1_A_1_, respectively. The association between allele dosage and phenotype is then estimated by the regression model:

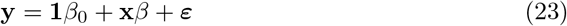

where **y** is an *n*-vector of phenotypes for *n* individuals, **1** is an *n*-vector of ones, *β*_0_ is the intercept, **x** is an *n*-vector of genotypes, *β* is the slope and represents the expected change in phenotype associated with each additional copy of A_1_, and ***ε*** is an *n*-vector of residuals with standard assumptions 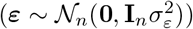. If *β* is significantly different from 0, there is an association between the genetic variant and the phenotype.

By connecting the GWAS regression model with the quantitative genetics theory described above, we can see that the GWAS coefficient *β* is a sample estimate of the allele substitution effect *α*. While the additive coding and linear regression strategy is statistically efficient and powerful for detecting associations, it constrains the fitted genotype effects to follow a linear relationship across the three genotype classes. When dominance is absent (*d ≠* 0), *α* and its estimate *β* can fully capture the relationship between genotype and phenotype. However, when *d* ≠ 0, *α* depends on the dominance effect and allele frequencies through the term *d*(*q* − *p*). Therefore, the *β* estimate may not accurately capture the actual relationship between genotype and phenotype. These properties of the *β* coefficient are directly relevant to post-GWAS applications such as phenotype prediction via polygenic scores (PGS), in which GWAS-estimated *β* coefficients are used.

PGS estimate an individual’s genetic value for a trait of interest by aggregating the effects of multiple genetic variants across the genome [7]. In a standard PGS, the score for individual *i* is calculated as the sum of allele dosages (*x*_*ij*_) weighted by their corresponding effect sizes 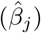:

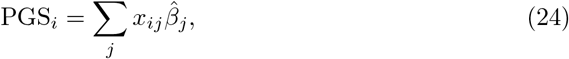

Many methods for PGS computation have been developed that make different assumptions about the distribution of genetic effects. The simplest methods to compute PGS use effect sizes estimated by GWAS and a procedure like clumping and thresholding (C+T) to select only independent variants associated with the trait [8–10]. More recently, multiple regression approaches have been developed for PGS computation, which adjust the GWAS-estimated effect sizes for linkage disequilibrium (LD) among variants [11–15]. However, the peculiarity of all these methods is that they use estimates of the allele substitution effects and constrain the PGS of the heterozygous genotype to be exactly in the middle of the PGS of the homozygous genotypes at each locus affecting the trait. Thus, all currently available PGS methods estimate the additive genetic values, depend on allele frequencies, and do not allow for dominance to be part of the genetic architecture of complex traits. If dominance is present, using an additive model may not be an important problem if one wants to predict *across generations* (as is the case for agricultural breeding), since dominance effects cannot be transmitted to the next generation (because genotypes segregate at meiosis and only alleles are transmitted). However, using an additive model could be a problem if the goal is individual predictions *within a generation* (as is the case for precision medicine), and dominance plays a role in the genetic architecture of the trait. Thus, PGS methods that are based on estimates of the average substitution effect are expected to be accurate only when there is additive gene action at all loci (*i.e., α* = *a*).

The role and importance of non-additive genetic effects for complex traits has been controversial. In this work, we only focus on dominance. Studies in model organisms and agricultural species have provided evidence for the importance of dominance for complex traits [16–18]. On the other hand, studies in humans have provided weak evidence for a role of dominance on complex traits [19–22]. While there may be many reasons for the apparent discrepancy between the results in humans and other species (*e.g*., human datasets are observational and noisy), it is fundamental to realize that most of the evidence for the limited importance of dominance to complex traits comes from variance component analysis. In particular, most studies estimated the proportion of phenotypic variance explained by dominance deviations (or dominance *variance*) and found it to be negligible compared to additive variance [19, 20, 22]. However, while variance component analysis is more powerful than single locus mapping, the lack of 1 - to-1 relationship between effects and variance components (*i.e*., dominance *effect* contributes to both additive and dominance *variance* components) highlighted by the quantitative genetics theory makes variance component analysis unsuitable to establish the importance of dominance effects on complex traits [23]. On the other hand, recent studies aimed at mapping loci with significant dominance effects have suggested that dominance may contribute to the genetic architecture of at least some human complex traits [21, 22].

A small number of studies have investigated whether including dominance can improve PGS performance. In livestock genomic prediction, Liu et al. incorporated locus-specific degree of dominance into the construction of the genomic relationship matrix in a GBLUP-type model and showed improved prediction accuracy for chicken and pig traits [24]. In humans, Pozarickij et al. constructed a PGS for refractive error that accounted for dominance and found that it slightly increased prediction accuracy [25]. Lu et al. showed that adding dominance terms for loci associated with variation in polygenic score prediction errors could improve PGS predictions for selected UK Biobank biomarkers [26]. More recently, GenBoost showed that accounting for dominance in the MHC locus could improve prediction for autoimmune diseases [27]. Another method, AB-PRS, further showed that fine-tuning a pre-trained PGS with effects not captured by the original score, including non-additive effects, could improve prediction for some complex traits and diseases [28].

However, existing approaches often incorporate dominance as additional selected terms, which increases the complexity of the model and may result in loss of power. Here, we developed PGS_*a,d*_, a simple polygenic scoring framework that incorporates both additive and dominance effects directly. Instead of relying only on allele dosage weighted by *β*, PGS_*a,d*_ assigns genotype-specific contributions based on estimates of *a* and *d*, which allows the heterozygote effect to deviate from the midpoint between the homozygotes and removes the dependency on allele frequencies. We tested PGS_*a,d*_ in simulations and then applied it to height, BMI, and 16 blood cell traits in the UK Biobank [29]. The results showed that PGS_*a,d*_ is competitive with the standard PGS for all traits, while outperforming it for some traits.

## Materials and methods

### Estimation and inference of the additive, dominance and average substitution effects

Following the classic quantitative genetic model described above and letting 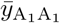, 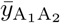, and 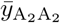 denote the mean phenotype values for genotypes A_1_A_1_, A_1_A_2_, and A_2_A_2_, respectively, the additive (*a*) and dominance (*d*) effects can be estimated as

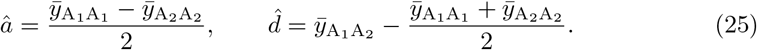

The standard errors of these estimates are:

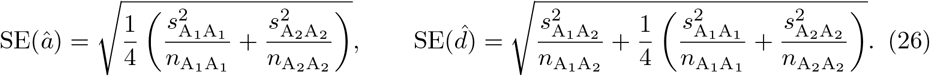

where 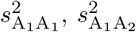, and 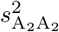 are the phenotypic variances for genotypes A_1_A_1_, A_1_A_2_, and A_2_A_2_, respectively.

We can test whether the estimates of the additive and dominance effects are significantly different from 0 using the following test statistics:

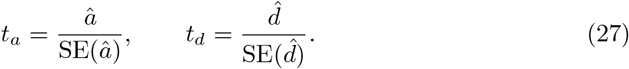

It can be shown that these test statistics follow *t* distributions with degrees of freedom computed using the Welch–Satterthwaite formula [30]. Expressions and detailed derivations are provided in S1 Text and S1 Fig.

As discussed in the Introduction, the average substitution effect (*α*) can be estimated and tested for significance via linear regression in GWAS.

### Computation of a new PGS method (PGS_a,d_)

Based on the theory, we developed a new PGS (PGS_*a,d*_) that incorporates genotype-dependent contributions from the additive and dominance effects for the set of variants affecting the trait. For individual *i*, the contribution of variant *j* is defined as:

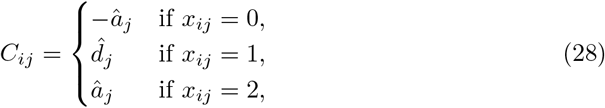

where *x*_*ij*_ is the genotype coded as the number of copies of the alternative allele. PGS_*a,d*_ is then defined as:

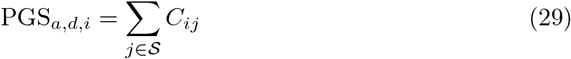

where S represents the set of variants affecting the trait, selected in some way.

In contrast, the conventional approach constructs PGS_*β*_ using the estimated average substitution effects obtained from GWAS. For individual *i*, the contribution of variant *j* is modeled as a linear function of genotype dosage:

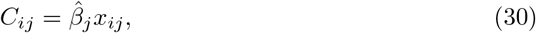

where *x*_*ij*_ ∈ {0,1,2} denotes the number of copies of the alternative allele, and 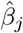 is the estimated effect size from GWAS. Thus, the polygenic score is:

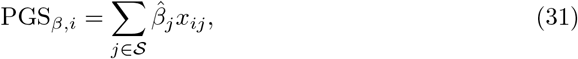

where S denotes the set of variants affecting the trait. Unlike PGS_*a,d*_, which assigns genotype-specific contributions based on â_*j*_ and 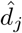, PGS_*β*_ assumes a strictly additive relationship between genotype and phenotype.

### Simulation study using UK Biobank genotypes

#### Data preparation

We conducted simulations using real genotypes from the UK Biobank array data [29]. Samples were restricted to unrelated White British individuals that did not exhibit discordance between genetic and self-reported sex, and were not identified as heterozygosity or genotype missingness outliers. A total of 337,467 individuals were retained, among which we randomly selected 200,000 individuals for the subsequent analyses. We applied variant-level quality control to the 200,000 sample dataset to exclude rare and low-quality variants, including filters on minor allele frequency (MAF *>* 0.01), minor allele count (MAC *>* 5), genotype missingness (missing rate *<* 0.1), and deviation from Hardy–Weinberg equilibrium (*P >* 1 × 10^−5^) using PLINK v.2.0 [31].

After quality control, 565,102 genetic variants were retained for downstream simulations. Finally, we randomly partitioned the sample set into a training set (180,000 individuals) and an independent test set (20,000 individuals).

#### Simulation of phenotypes

To evaluate performance under different patterns of gene action, we performed simulations under three genetic architectures: (i) purely additive; (ii) combined additive–dominant; and (iii) purely dominant. In all settings, additive effects were assigned to 1,000 randomly sampled causal loci, with *a*_*j*_ ∼ *N* (0, 2). Under the purely additive architecture, dominance effects were fixed to zero (*d*_*j*_ = 0 ∀ *j*). Under the combined additive–dominant architecture, dominance effects were assigned to 500 out of 1000 causal loci. For each of these loci, we either assigned complete dominance (*d*_*j*_ = ±*a*_*j*_) or partial dominance (*d*_*j*_ = ±*u*_*j*_*a*_*j*_, where *u*_*j*_ ∼ *u*(0.2, 0.8)) with equal probability. The sign of the dominance effect was also assigned with equal probability in both cases. Under the purely dominant architecture, dominance effects were assigned to all 1,000 loci using the same procedure as in the additive–dominant architecture.

Given the assigned additive and dominance effects, we generated phenotypes using the 1,000 causal loci with an independent environmental noise. For causal variant *j* with effects *a*_*j*_ and *d*_*j*_, the per-variant genotypic contribution for individual *i* was defined as:

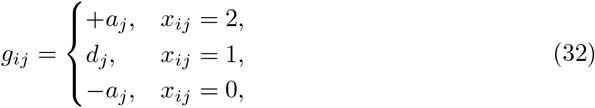

where *x*_*ij*_ denotes the genotype coded as the number of copies of the alternative allele carried by individual *i* at variant *j*. The simulated phenotype for individual *i* was generated as:

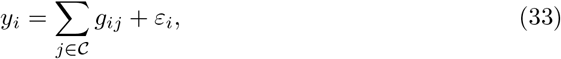

where denotes the set of causal loci and 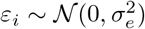 is a random error term. The residual variance 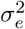 was computed such that the genomic heritability was 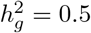.

We generated 20 replicates for each genetic architecture by resampling causal variants, effect sizes, and error, while keeping the genotype matrix and sample partition fixed.

#### Statistical analysis

We performed genome-wide association analysis (GWAS) on the training set to obtain estimates of the effect size 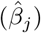 and the corresponding *P* -value for each variant *j* using PLINK v2.0. We selected variants based on *P* -value thresholds of 10^−3^, 10^−5^, 10^−8^ and subsequently clumped selected variants based on linkage disequilibrium (LD) using *r*^2^ thresholds of 0.1, 0.3, and 0.5 (`--clump-r2`) within a 1,000 kb window (`--clump-kb 1000`). We then constructed polygenic scores for all nine combinations of *P* -value and LD thresholds, and selected the combination yielding the best prediction performance in the test set.

We calculated the traditional PGS_*β*_ for individual *i* as:

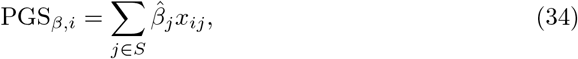

where *x*_*ij*_ ∈ { 0, 1, 2} represents the number of alternative allele copies at variant *j* and *S* is the set of variants retained after clumping and thresholding (C+T).

To compute PGS_*a,d*_, â and 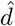 were obtained for the set *S* using eq. 25. 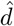 was tested for significance using *t*_*d*_ in eq. 27 and a two-sided *P* -value. PGS_*a,d*_ for individual *i* was calculated as:

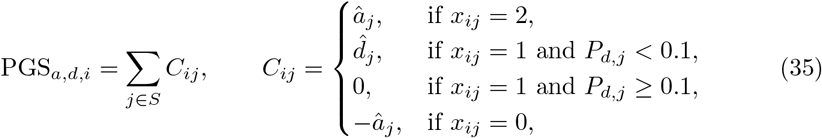

We evaluated prediction performance in the test set by comparing observed phenotypes (**y**) with the corresponding PGSs in terms of prediction *R*^2^ and root mean squared error (RMSE). *R*^2^ was computed from the linear regression of **y** on the PGS, while RMSE was defined as:

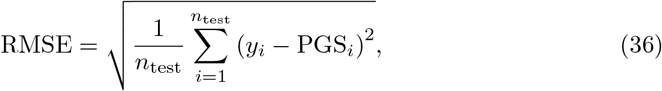

where *y*_*i*_ denotes the observed phenotype of individual *i*, PGS_*i*_ the corresponding polygenic score, and *n*_test_ the number of individuals in the test set.

### Application to real traits in the UK Biobank

#### Data preparation

To evaluate our method in a real data application, we applied the same analytical framework to 16 blood cell traits from the UK Biobank (Table 1).

**Table 1.**
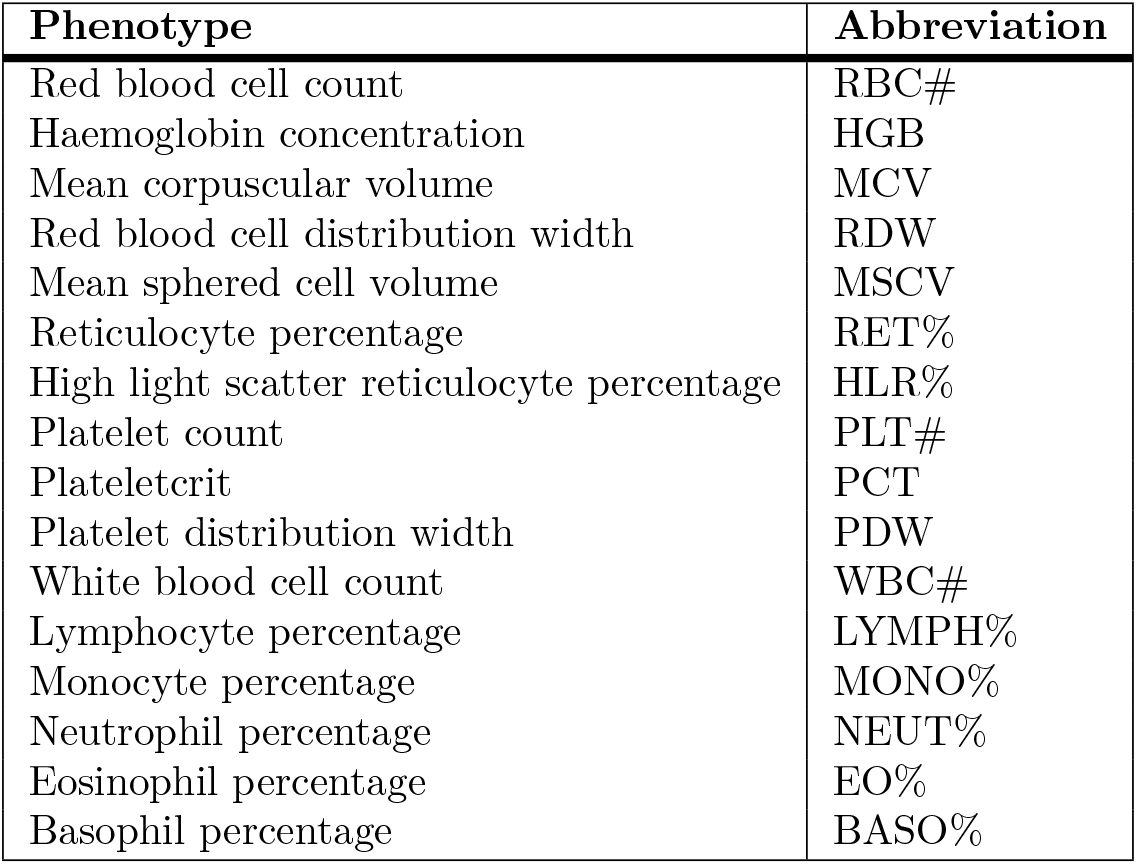
List of the 16 blood cell traits and their abbreviations.

We implemented similar sample control procedures to the simulations, including restriction to unrelated White British individuals and exclusion of individuals with discordance between genetic and self-reported sex, as well as heterozygosity or genotype missingness outliers. We also excluded pregnant individuals at baseline assessment. To reduce potential confounding from pre-existing conditions affecting hematological measurements, we further excluded individuals with hospital in-patient records indicating blood-related diseases using ICD10 according to the disease exclusion criteria described by [32]. We retained only individuals with complete measurements for all 16 blood cell traits. After applying exclusion criteria, 273,795 participants remained. Following [32], we normalized each of the 16 blood cell traits using quantile normalization to approximate a standard normal distribution. Multivariate outlier detection was subsequently performed using a Mahalanobis distance-based approach. For each individual *i*, we computed the squared Mahalanobis distance 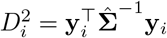, where **y**_*i*_ denotes the 16-dimensional vector of normalized trait values for individual *i*, and 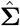 is the sample covariance matrix of the normalized traits. We excluded individuals with squared Mahalanobis distance exceeding the 99^th^ percentile of a *χ*^2^ distribution with 16 degrees of freedom, resulting in a final sample of 265,131 participants.

We performed genotype filtering on the 265,131 participants using the same quality control criteria as in the simulation analyses, including filters on minor allele frequency (MAF *>* 0.01), minor allele count (MAC *>* 5), genotype missingness (missing rate *<* 0.1), and deviation from Hardy–Weinberg equilibrium (*P >* 1 × 10^−5^). After quality control, 565,361 genetic variants were retained for downstream analyses.

To assess the generalizability of our framework, we also selected height and body mass index (BMI), two complex traits with predominantly additive and highly polygenic architectures [33–35]. Starting from the sample used for the blood cell traits analysis, we excluded individuals missing height or BMI measurements. In total, we retained 264,663 individuals for height and 264,432 individuals for BMI.

#### Statistical analysis

##### Detection of loci exhibiting dominance

Before evaluating prediction performance, we performed GWASs for all traits using the full sample. GWAS was conducted using LDAK-KVIK [36], adjusting for sex, age, age^2^, age-by-sex interaction, and the first 10 genetic principal components (PCs). Age and PCs were standardized prior to model fitting.

Based on GWASs results, variants reaching genome-wide significance (*P <* 5 × 10^−8^; `--clump-p1 5e-8`) were subjected to LD clumping using an *r*^2^ threshold of 0.1 (`--clump-r2 0.1`) within a 1,000 kb window (`--clump-kb 1000`) to obtain an approximately independent set of lead variants. We then estimated the dominance effect for this clumped set of variants, and tested for significance. Significant variants were identified using a Bonferroni-corrected *P* -value threshold of 0.05*/*(*N*_SNPs_ = 78, 310), where *N*_SNPs_ represents the total number of variants tested across 18 traits, including 16 blood cell traits, height, and BMI. To remove potential confounding effects of covariates, we regressed the phenotypes on the same set of covariates as in the GWAS and used the resulting residuals for the estimation of the dominance effects.

##### Computation and evaluation of PGSs

To evaluate the prediction performance of the polygenic scores, we used 5-fold cross-validation. We randomly partitioned the sample into five non-overlapping groups of approximately equal size. At each iteration, four groups were used as the training set and the remaining group was held out as the test set. Within each iteration, we applied the same analytical framework described above for the simulations to the training set, including GWAS using LDAK-KVIK, genome-wide significant variant selection (*P <* 1 × 10^−8^), LD clumping (`--clump-p1 1e-8, --clump-r2 0.1, --clump-kb 1000`), and estimation of additive and dominance effects. We chose the genome-wide significant threshold and LD clumping parameters based on the best-performing combination identified in the simulation analysis. We computed PGS_*β*_ and PGS_*a,d*_ in the test set using eq. 34 and eq. 35, respectively. We evaluated prediction accuracy by comparing the PGS with the true phenotypes (adjusted for the same covariates used in the GWAS using linear regression) using prediction *R*^2^ and RMSE. For each trait, we compared these metrics between PGS_*β*_ and PGS_*a,d*_ using a paired *t* −test and a significance threshold of 0.05*/*18 = 2.78 × 10^−3^ to account for multiple testing.

## Results

### Simulation study using UK Biobank genotypes

We evaluated the performance of PGS_*a,d*_ and PGS_*β*_ across the simulated additive, combined additive and dominant, and dominant genetic architectures. Across all simulation settings, the relative performance of the two methods varied with the underlying genetic architecture (Figure 3).

**Fig 3.**
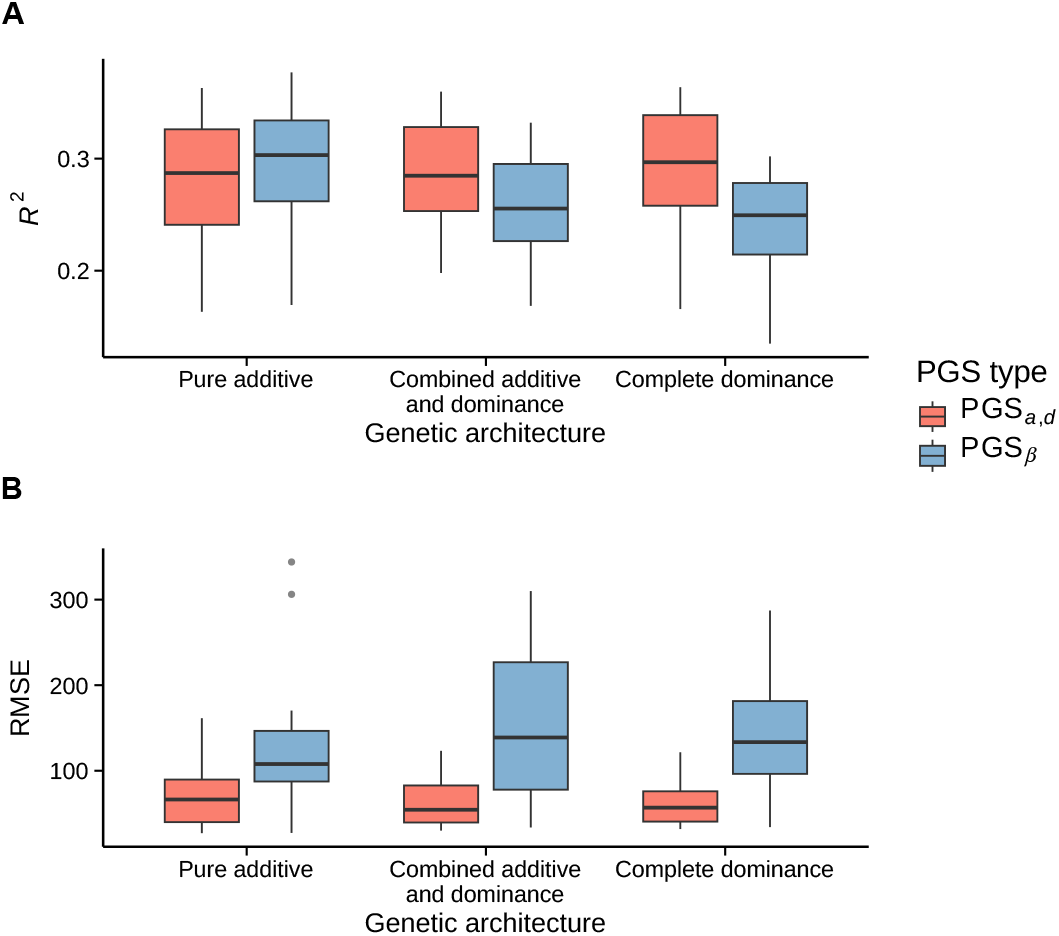
Comparison of prediction accuracy between PGS_*a,d*_ and PGS_*β*_ in simulations. Prediction performance was evaluated across three simulated genetic architectures, including purely additive, combined additive and dominance, and complete dominance. (A) Comparison of *R*^2^ across three genetic architectures. Higher *R*^2^ indicates better prediction accuracy. (B) Comparison of RMSE across three genetic architectures. Lower RMSE indicates lower prediction errors. Boxplots summarize performance across simulation replicates under each genetic architecture.

In general, while PGS_*a,d*_ and PGS_*β*_ showed similar performance under an additive genetic architecture, differences between the two approaches became apparent when dominance was part of the genetic architecture. Figure 3A shows the prediction *R*^2^ for the three simulated architectures using a clumping *P* -value threshold of *P <* 1 × 10^−8^ and LD threshold of *r*^2^ = 0.1. PGS_*a,d*_ achieved comparable *R*^2^ to PGS_*β*_ under the purely additive architecture, but showed higher *R*^2^ under both the additive–dominant and purely dominant architectures. The difference in *R*^2^ increased with the contribution of dominance effects. PGS_*a,d*_ consistently yielded lower RMSE than PGS_*β*_ across all genetic architectures, including the additive scenario (Figure 3B). The difference between the two methods increased with the contribution of dominance effects, with the largest difference observed under the dominant architecture. In addition, PGS_*β*_ showed greater variability in RMSE across replicates compared to PGS_*a,d*_.

These patterns were observed consistently across combinations of clumping *P* -value and LD thresholds (S2 Fig, S3 Fig), indicating that the relative performance of the methods was not sensitive to parameter choice. Among all the combinations tested, we chose *P <* 1 × 10^−8^ and *r*^2^ = 0.1 for subsequent analyses as a conservative and commonly used combination that provides a set of relatively independent variants with strong evidence of association for downstream analyses.

### Application to real traits in the UK Biobank

#### Detection of dominance effects

After quality control, we obtained a total of 78,310 variants for analysis across 18 traits. In an effort to understand whether dominance played a role in the genetic architecture of the blood cell traits, we performed GWAS for all blood cell traits and scanned for non-zero dominance effects of the clumped lead GWAS-significant variants. We identified genome-wide significant loci for the majority of traits using a Bonferroni-corrected threshold of 0.05/78,310 = 6.38 × 10^−7^ as shown in Table 2 and S1 Table.

**Table 2.** Genome-wide significant loci exhibiting dominance effects across blood cell traits. Peak width denotes the genomic span of the associated locus, and No. SNPs indicates the number of significant variants within that locus.

| Trait | Lead variant | Chr | *P*-value | Peak width (bp) | No. SNPs |
| --- | --- | --- | --- | --- | --- |
| RBC# | rs7775698 | 6 | $1.74 \times 10^{-11}$ | 21324 | 7 |
| RBC# | rs507666 | 9 | $4.62 \times 10^{-14}$ | 17103 | 8 |
| HGB | rs507666 | 9 | $6.28 \times 10^{-11}$ | 17103 | 8 |
| MCV | rs7775698 | 6 | $1.41 \times 10^{-17}$ | 24273 | 8 |
| MCV | rs4820268 | 22 | $1.10 \times 10^{-13}$ | 634 | 3 |
| RDW | rs3811444 | 1 | $7.91 \times 10^{-11}$ | 0 | 1 |
| RDW | rs445925 | 19 | $1.46 \times 10^{-16}$ | 0 | 1 |
| RDW | rs2413450 | 22 | $6.31 \times 10^{-11}$ | 634 | 3 |
| MSCV | Affx-52122172 | 1 | $1.27 \times 10^{-16}$ | 170415 | 5 |
| MSCV | rs857721 | 1 | $5.06 \times 10^{-12}$ | 95406 | 5 |
| MSCV | rs13194491 | 6 | $1.85 \times 10^{-8}$ | 0 | 1 |
| MSCV | rs7775698 | 6 | $1.09 \times 10^{-8}$ | 16589 | 6 |
| MSCV | rs16926246 | 10 | $4.13 \times 10^{-8}$ | 0 | 1 |
| RET% | Affx-52122172 | 1 | $1.61 \times 10^{-28}$ | 29196 | 2 |
| RET% | rs857721 | 1 | $2.59 \times 10^{-9}$ | 35439 | 4 |
| RET% | rs57467915 | 2 | $1.29 \times 10^{-10}$ | 84111 | 2 |
| RET% | rs16926246 | 10 | $1.51 \times 10^{-12}$ | 0 | 1 |
| HLR% | rs57467915 | 2 | $7.16 \times 10^{-11}$ | 84111 | 2 |
| HLR% | rs16926246 | 10 | $3.56 \times 10^{-10}$ | 0 | 1 |
| HLR% | rs445925 | 19 | $5.63 \times 10^{-10}$ | 0 | 1 |
| PLT# | rs139898146 | 1 | $6.43 \times 10^{-33}$ | 242892 | 11 |
| PLT# | rs74227709 | 1 | $2.87 \times 10^{-8}$ | 2819 | 2 |
| PLT# | rs1354034 | 3 | $6.47 \times 10^{-26}$ | 16027 | 2 |
| PLT# | rs7775698 | 6 | $2.42 \times 10^{-8}$ | 16589 | 6 |
| PLT# | rs342281 | 7 | $1.76 \times 10^{-7}$ | 13003 | 3 |
| PLT# | rs7961894 | 12 | $7.70 \times 10^{-15}$ | 0 | 1 |
| PLT# | rs11082304 | 18 | $8.81 \times 10^{-13}$ | 0 | 1 |
| PCT | rs2236055 | 1 | $1.17 \times 10^{-7}$ | 0 | 1 |
| PCT | rs139898146 | 1 | $1.50 \times 10^{-37}$ | 242892 | 11 |
| PCT | rs1354034 | 3 | $2.57 \times 10^{-8}$ | 0 | 1 |
| PCT | rs4895441 | 6 | $1.85 \times 10^{-8}$ | 16589 | 6 |
| PDW | Affx-52122172 | 1 | $4.35 \times 10^{-16}$ | 170415 | 5 |
| PDW | rs7961894 | 12 | $3.99 \times 10^{-21}$ | 0 | 1 |
| PDW | rs11082304 | 18 | $9.90 \times 10^{-10}$ | 0 | 1 |
| PDW | rs463312 | 20 | $3.65 \times 10^{-53}$ | 3286 | 2 |
| EO% | rs2394173 | 6 | $1.03 \times 10^{-9}$ | 206481 | 14 |
| EO% | rs4410767 | 6 | $6.24 \times 10^{-7}$ | 0 | 1 |
| EO% | rs34108746 | 11 | $1.09 \times 10^{-53}$ | 69405 | 2 |

In total, 141 significant variants were grouped into 38 genomic regions (S4 Fig). The number of genomic regions with significant dominance effects varied considerably across traits, ranging from 1 region for HGB to 7 regions for PLT#. The number of significant variants within each region also varied, from single-variant peaks to broader regions containing multiple nearby significant variants.

Platelet-related traits showed some of the strongest and most numerous signals, including 26 significant variants across seven regions for PLT# and 19 significant variants across four regions for PCT. Several red blood cell-related traits also showed multiple significant regions, including RDW, MSCV, and RET%. For example, the peak underlying *rs139898146* on chromosome 1 comprised 11 significant variants for both PCT and PLT#, indicating a broad signal detected by the analysis. Several traits, including MSCV, PLT#, and PDW, showed significant peaks at multiple genomic regions, suggesting that dominance effects for these traits were not driven by a single locus.

Some signals were particularly strong, such as *rs34108746* on chromosome 11 with EO% (*P* = 1.09 × 10^−53^), representing one of the most significant dominance signals identified. Variants near *rs507666* on chromosome 9 showed significant dominance effects for both HGB and RBC#, consistent with a shared genetic basis across red blood cell traits. In addition, *rs7775698* (MCV, RBC#, and PLT#) and *rs857721* (MSCV and RET%) were associated with multiple traits, suggesting potential pleiotropic dominance effects. In contrast, no genome-wide significant dominance loci were identified for BASO%, MONO%, NEUT%, or LYMPH% after Bonferroni correction.

#### Evaluation of PGS prediction accuracy

We compared the predictive performance of PGS_*β*_ and PGS_*a,d*_ across all blood cell traits using both *R*^2^ and RMSE. Overall, PGS_*a,d*_ and PGS_*β*_ performed comparably in terms of *R*^2^ (Figure 4, S2 Table), even though PGS_*β*_ tended to achieve slightly higher *R*^2^. On the other hand, PGS_*a,d*_ resulted in lower RMSE than PGS_*β*_ (Figure 4, S3 Table) for 13 out of 16 traits, with a statistically significant difference observed for RDW, MONO%, NEU%, and RET% after strict Bonferroni correction accounting for all the traits (0.05*/*18 = 2.78 ×10^−3^). The reduction in RMSE using PGS_*a,d*_ was particularly notable for RDW and MONO%.

**Fig 4.**
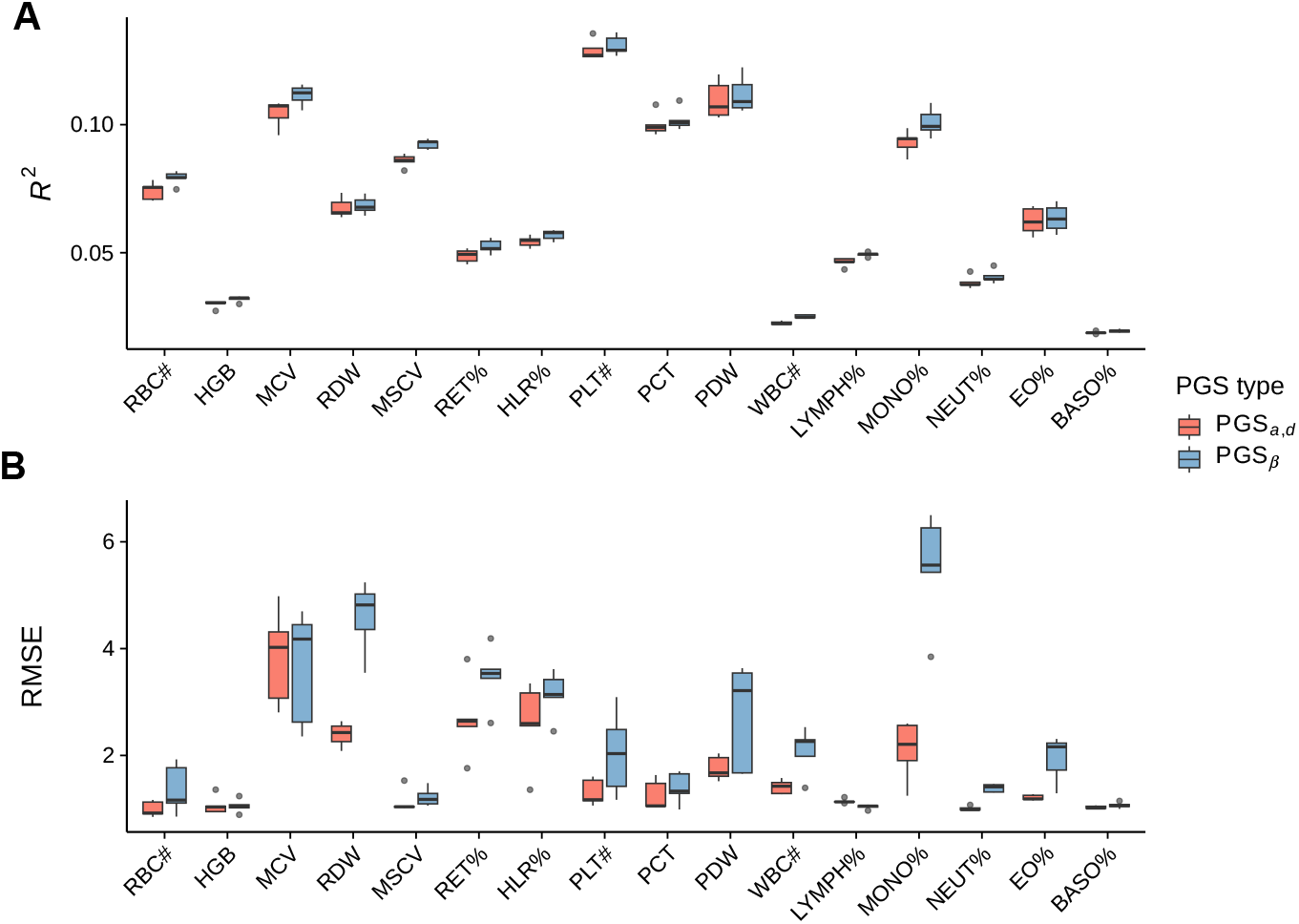
Comparison of prediction accuracy between PGS_*a,d*_ and PGS_*β*_ across 16 blood cell traits using 5-fold cross-validation. (A) Comparison of coefficient of determination (*R*^2^) across 16 blood cell traits. Higher *R*^2^ indicates better prediction accuracy. (B) Comparison of root mean squared error (RMSE) across 16 blood cell traits. Lower RMSE indicates lower prediction errors. For each trait, boxplots summarize performance across the 5 cross-validation folds.

We further evaluated the performance of PGS_*β*_ and PGS_*a,d*_ for two additional complex traits, height and BMI, that are thought to have a mostly additive genetic architecture. In contrast to blood cell traits, Manhattan plots of dominance effects revealed limited evidence of genome-wide significant loci for either trait after multiple testing correction (S4 Fig), suggesting that dominance effects are generally weak or null for these traits. Consistent with the absence of strong dominance effects, PGS_*β*_ and PGS_*a,d*_ achieved very similar *R*^2^ for both height and BMI (Figure 5, S2 Table). In terms of RMSE, PGS_*a,d*_ resulted in significantly lower value than PGS_*β*_ for height after Bonferroni correction and very similar value for BMI (S3 Table).

**Fig 5.**
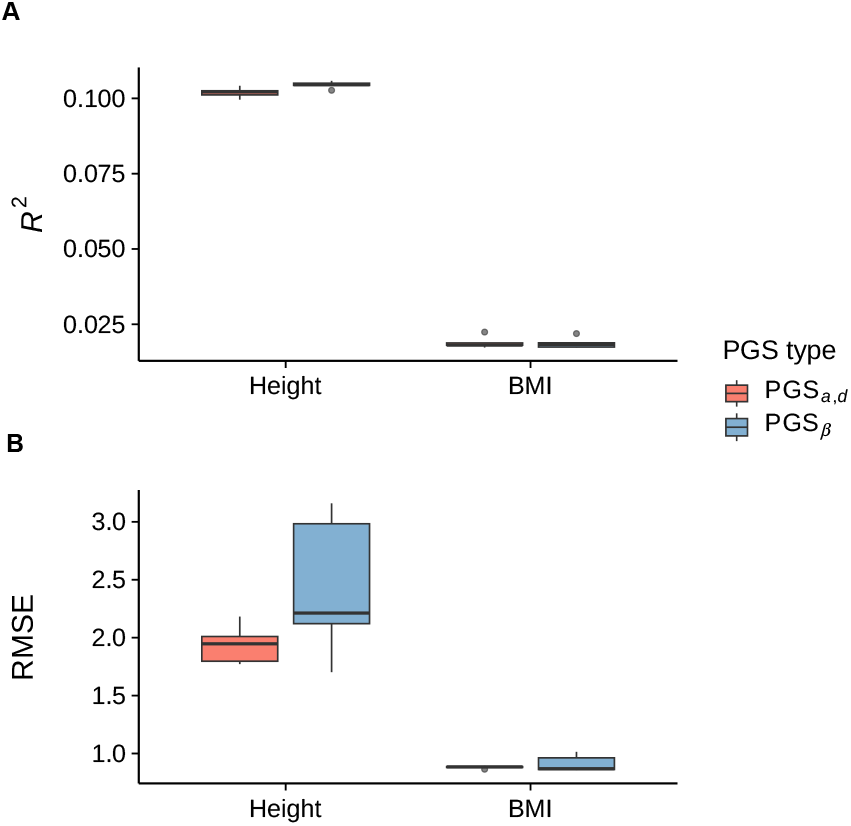
Comparison of prediction accuracy between PGS_*a,d*_ and PGS_*β*_ for height and BMI. (A) Comparison of coefficient of determination (*R*^2^). Higher *R*^2^ indicates better prediction accuracy. (B) Comparison of root mean squared error (RMSE). Lower RMSE indicates lower prediction errors. For each trait, boxplots summarize performance across the 5 cross-validation folds.

We then sought to understand why some traits showed markedly lower RMSE when using PGS_*a,d*_, whereas some other traits did not. The magnitude of RMSE improvement appeared to be related to the size of estimated dominance effects – traits with larger mean absolute dominance effect sizes tended to show greater RMSE reduction from PGS_*a,d*_ relative to PGS_*β*_ (Spearman *ρ* = 0.496, *P* = 0.036; Figure 6).

**Fig 6.**
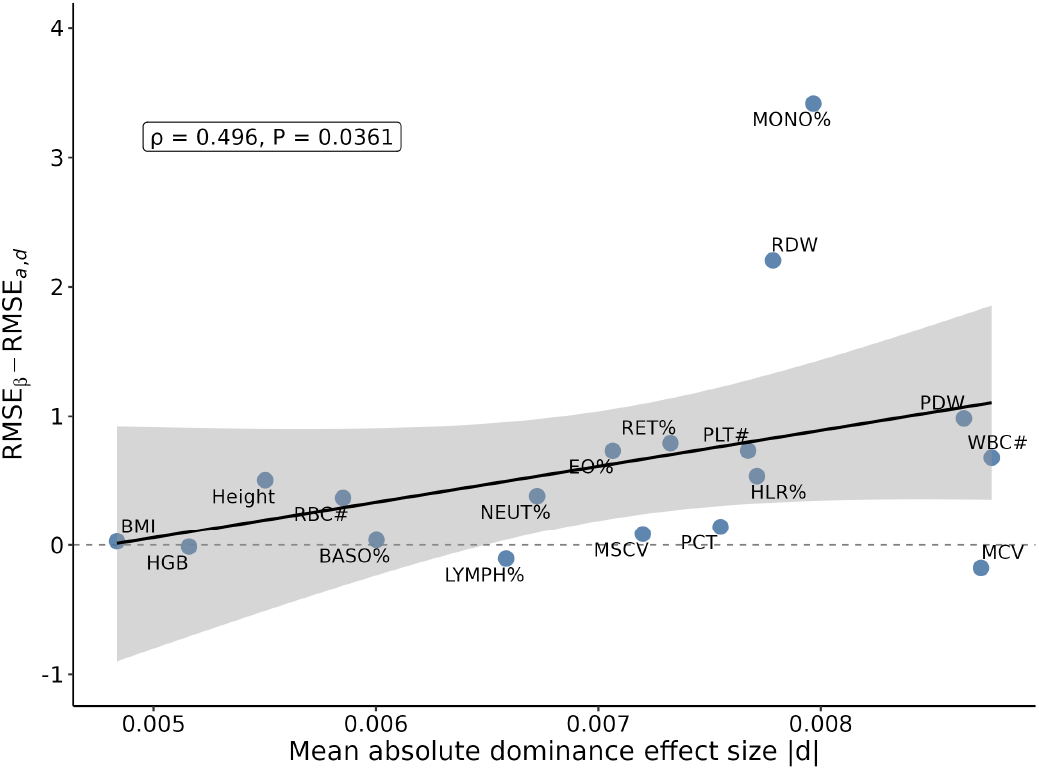
Relationship between mean absolute dominance effect size and RMSE difference between PGS_*β*_ and PGS_*a,d*_ across traits. Each point represents one trait. The x-axis shows the mean absolute dominance effect size and the y-axis shows the mean RMSE improvement, defined as RMSE_*β*_ minus RMSE_*a,d*_, where positive values indicate lower prediction error for PGS_*a,d*_ compared with PGS_*β*_. The fitted line shows the linear trend, with the shaded area representing the 95% confidence interval. Spearman correlation statistics are shown in the plot.

## Discussion

In this study, we developed PGS_*a,d*_, a new polygenic score framework that directly uses estimated genotypic effects (additive and dominance effects) rather than relying on allele dosage weighted by GWAS-estimated allele substitution effect. The simulation and real trait analyses support the potential value of PGS_*a,d*_ for individual-level predictions, particularly when dominance contributes to the genetic architecture. Across simulations, PGS_*a,d*_ showed comparable performance to the commonly-adopted PGS based on allele substitution effect (PGS_*β*_) when the genetic architecture was purely additive. As the contribution of dominance effects increased, the advantages of PGS_*a,d*_ became more pronounced, with higher correlation (*R*^2^) and lower error (RMSE) with the true phenotypes. This pattern is consistent with the distinction between allele substitution effects and genotypic effects. As shown above, *β* is an estimator of the allele substitution effect, which measures the effect of an allele on a phenotype *in the next generation*. However, the predictions obtained using allele substitution effects (*i.e*., the additive genetic value) do not match the actual genotypic value when dominance is present. Consequently, PGS_*β*_ provides lower accuracy in this scenario. By contrast, PGS_*a,d*_ can retain deviations of the heterozygote from the additive expectations and more closely align the score to the actual genotypic value for *within-generation* prediction.

In the real traits application, PGS_*a,d*_ had significantly lower RMSE for RDW, MONO%, NEUT%, and RET% after strict multiple testing correction. However, PGS_*a,d*_ provided similar or slightly lower *R*^2^ than PGS_*β*_. This is not surprising since *R*^2^ reflects how well a score captures phenotypic variation in the population, which theory predicts to be mostly additive even when gene action is not additve [37]. In addition, our simulations showed *R*^2^ to be slightly higher for PGS_*β*_ than PGS_*a,d*_ for completely additive architectures. On the other hand, RMSE measures the discrepancy between predicted and observed phenotypes and is therefore a better measure of individual-level prediction error. In agreement with simulations, traits with larger estimated dominance effects generally showed greater RMSE improvement of PGS_*a,d*_ over PGS_*β*_. For height and BMI, two highly polygenic traits with predominantly additive architectures [33–35], PGS_*a,d*_ still performed similarly to PGS_*β*_ while maintaining slightly lower RMSE. Together, these results suggest that modeling additive and dominance effects directly is a robust strategy for polygenic score computation.

A main advantage of PGS_*a,d*_ is that the genotypic effects, *a* and *d*, do not depend on allele frequency. In contrast, the average allele substitution effect underlying PGS_*β*_ is frequency-dependent when dominance is present. Previous approaches incorporating dominance have generally extended an additive PGS by introducing additional terms to capture deviations from the additive expectation (*i.e*., dominance deviations in quantitative genetics terminology). For example, Pozarickij et al. added a heterozygote-specific deviation term for variants showing complete dominance or recessive patterns [25], while Lu et al. incorporated regression coefficients for indicator variables representing dominant or recessive genotype patterns into a conventional PGS constructed from GWAS-estimated effects [26]. More recently, AB-PRS was introduced as a way to fine-tune a pre-trained PGS by capturing residual SNP effects, including non-additive effects and other uncaptured predictive signals, after conditioning on the original score. This approach was shown to improve predictive performance for several complex traits and diseases [28]. However, because these approaches incorporate allele substitution effects, differences in allele frequencies between discovery and target populations may affect the estimated genetic effects and their transferability. The idea of allowing heterozygote effects to deviate from the additive midpoint has also been explored in genomic prediction in livestock. For example, Liu et al. parameterized this deviation through locus-specific degree of dominance, rather than directly through the genotypic effects, and used this parameterization to compute a genomic relationship matrix in GBLUP-type model [24]. This approach assumes an infinitesimal architecture of every trait and remains dependent on allele frequencies. PGS_*a,d*_ instead provides a formulation based directly on genotypic effects that are independent of allele frequency and may be more robust across populations. However, the present analyses were restricted to White British participants from UK Biobank. Thus, future work will need to evaluate the robustness of PGS_*a,d*_ in large diverse populations.

Importantly, PGS_*a,d*_ should, at this stage, be viewed as a proof-of-concept for evaluating whether explicitly modeling genotypic effects directly can improve polygenic prediction accuracy. In the current implementation, we applied the clumping-and-thresholding (“C+T”) strategy to restrict the score to relatively independent variants with evidence of association with the trait, thereby reducing redundancy among correlated variants. While C+T is easy to implement, it is well-known that methods that estimate the effects of multiple genetic variants jointly (accounting for LD) usually achieve higher prediction accuracy than C+T [13, 14]. In fact, current state-of-the-art methods produced higher accuracy for the same blood cell traits analyzed here [38]. However, the goal of our work was not to develop the most accurate PGS method – it was to evaluate whether modeling gene action could be useful for polygenic prediction. Our “like-with-like” comparison with PGS_*β*_ showed that modeling genotypic effects is a promising approach and is worth pursuing further with the development of more accurate prediction methods.

In addition to computing PGS_*a,d*_, our genome-wide scan for significant dominance effects provided additional insight into the role of dominance in the genetic architecture of the traits analyzed. Among the 16 blood cell traits, significant dominant effects were identified for RBC#, HGB, MCV, RDW, MSCV, RET%, HLR%, PLT#, PCT, PDW, and EO%. We compared our findings with those reported by Palmer *et al*., who conducted a large-scale systematic investigation of dominance across more than 1,000 phenotypes in the UK Biobank [22]. It is important to note that our study and that of Palmer *et al*. differ in the parameterization of dominance. For each genetic variant, they tested for significant evidence of dominance using an allele frequency-dependent encoding for the three genotypes [22]. This particular encoding was developed for the goal of variance decomposition, because it results in estimates of the dominance variance that are orthogonal to estimates of the additive variance [19]. However, our goal was not estimating variance but mapping variants showing non-zero dominance effect; therefore, we developed a statistical framework to test for that hypothesis directly.

Several variants and genomic regions with significant dominance effects overlapped between our study and that of Palmer *et al*.. Both studies identified the *RHD* locus across multiple hematological phenotypes. Palmer *et al*. reported significant dominance at this locus for 10 blood cell traits, while we identified variants with significant dominance effects in the same region for MSCV, PCT, PDW, PLT#, and RET%. We also observed overlap at the *ARHGEF3* locus, where Palmer *et al*. identified *rs1354034* for PDW, while we identified the same variant for PCT and PLT#. In addition, both studies identified the missense variant *rs34108746* in *PRG3* for EO% and *rs4820268* in *TMPRSS6* for MCV. In addition to these overlapping regions, we identified significant dominance effects at several loci that were not highlighted by Palmer *et al*.. For example, we observed a dominance signal at the *HBS1L-MYB* locus, represented by *rs7775698*, across MCV, RBC#, MSCV and PLT#. This variant may be involved in regulatory processes of blood cell traits as genetic variation at the *HBS1L-MYB* locus has been shown to influence erythroid progenitor proliferation, RBC#, and MCV [39–41]. We also identified significant dominance effects at the *ABO* locus, which aligns with recent evidence for non-additive effects on plasma protein abundance, including dominance and epistatic effects involving *ABO* variants [42].

## Data Availability

The genotype and phenotype data used in our analyses are available from UK Biobank (https://www.ukbiobank.ac.uk/).

## Supporting information

**S1 Fig. Q-Q plots of the test statistics for estimated additive and dominance effects under the null hypothesis**. The empirical distributions of the test statistics are compared with the theoretical *t*-distribution quantiles for (A) the estimated additive effect (â) and (B) the estimated dominance effect 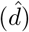. The test statistics closely follow the theoretical quantiles and align along the diagonal, indicating that the variance estimators and corresponding *t*-statistics are well calibrated under the null hypotheses *a* = 0 and *d* = 0.

**S2 Fig. Comparison of** *R*^2^ **between PGS**_*a,d*_ **and PGS**_*β*_ **across parameter combinations and genetic architectures**. Prediction accuracy was evaluated using *R*^2^ across combinations of clumping *P* -value thresholds (*P <* 10^−3^, 10^−5^, and 10^−8^) and LD *r*^2^ thresholds (*r*^2^ = 0.1, 0.3, and 0.5) under three simulated genetic architectures. (A) Additive architecture. (B) Combined additive and dominant architecture. (C) Dominant architecture. Model performance was summarized across simulation replicates for each parameter combination.

**S3 Fig. Comparison of RMSE between PGS**_*a,d*_ **and PGS**_*β*_ **across parameter combinations and genetic architectures**. Prediction accuracy was evaluated using the root mean squared error (RMSE) across combinations of clumping *P* -value thresholds (*P <* 10^−3^, 10^−5^, and 10^−8^) and LD *r*^2^ thresholds (*r*^2^ = 0.1, 0.3, and 0.5) under three simulated genetic architectures. (A) Additive architecture. (B) Combined additive and dominant architecture. (C) Dominant architecture. Model performance was summarized across simulation replicates for each parameter combination.

**S4 Fig. Manhattan plots of dominance effects**. Manhattan plots for dominance effects 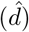 for variants analyzed across blood cell traits, height, and BMI. Each panel is a different trait and the dashed lines indicate the Bonferroni-corrected significance thresholds.

**S1 Text. Supplementary Notes**. Expressions and derivations for the test statistics of estimated additive and dominance effects.

**S1 Table. Loci with genome-wide significant dominance effect**. This table summarizes significant dominance signals identified across 16 blood cell traits, height, and BMI after strict Bonferroni corrections. For each genomic region, the table reports the trait, lead SNP, chromosome, lead SNP position, *P* value for dominance effect, peak boundaries, peak width, number of significant SNPs within the peak, additional significant SNPs assigned to the same peak, and the number of tested SNPs for each trait.

**S2 Table. Paired comparison of** *R*^2^ **between PGS**_*a,d*_ **and PGS**_*β*_ **across all traits**. This table summarizes paired t-test results comparing *R*^2^ between PGS_*a,d*_ and PGS_*β*_ across 5-fold cross-validation test sets for each of the 16 blood cell traits, height, and BMI. For each trait, the table reports the mean *R*^2^ for each method, the mean difference between methods, the *t* statistic, the *P* -value, and significance after Bonferroni correction.

**S3 Table. Paired comparison of RMSE between PGS**_*a,d*_ **and PGS**_*β*_ **across all traits**. This table summarizes paired *t* -test results comparing RMSE between PGS_*a,d*_ and PGS_*β*_ across 5-fold cross-validation test sets for each of the 16 blood cell traits, height, and BMI. For each trait, the table reports the mean RMSE for each method, the mean difference between methods, the *t* statistic, the *P* -value, and significance after Bonferroni correction.

## Data and code availability

The genotype and phenotype data used in our analyses are available from UK Biobank (https://www.ukbiobank.ac.uk/). Code used in the analyses is available at https://github.com/morgantelab/prs_effects.

## Acknowledgments

This research was conducted using the UK Biobank Resource under application number 129216. Research reported in this publication was supported by the National Institute of General Medical Sciences of the National Institutes of Health under Award Number R35GM146868 to FM. The content is solely the responsibility of the authors and does not necessarily represent the official views of the National Institutes of Health.

## Authors contributions

Conceptualization: F.M., T.F.C.M; Data curation: Y.S., F.M.; Formal analysis: Y.S.; Funding acquisition: F.M.; Investigation: Y.S., F.M., T.F.C.M.; Methodology: Y.S., F.M., T.F.C.M.; Project administration: F.M., T.F.C.M.; Resources: F.M., T.F.C.M.; Software: Y.S.; Supervision: F.M., T.F.C.M.; Validation: Y.S., F.M., T.F.C.M.; Visualization: Y.S.; Writing–original draft: Y.S.; Writing–review & editing: Y.S., F.M., T.F.C.M.

## References

1. Mackay TFC, Stone EA, Ayroles JF. The genetics of quantitative traits: challenges and prospects. Nature Reviews Genetics. 2009;10(8):565–77. Available from: https://doi.org/10.1038/nrg2612. doi:10.1038/nrg2612.

2. Goddard ME, Kemper KE, MacLeod IM, Chamberlain AJ, Hayes BJ. Genetics of complex traits: prediction of phenotype, identification of causal polymorphisms and genetic architecture. Proceedings of the Royal Society B: Biological Sciences. 2016;283(1835):20160569. Available from: https://doi.org/10.1098/rspb.2016.0569. doi:10.1098/rspb.2016.0569.

3. Fisher RA. XV. —The correlation between relatives on the supposition of Mendelian inheritance. Earth and Environmental Science Transactions of the Royal Society of Edinburgh. 1919;52(2):399–433.

4. Falconer DS, Mackay TFC. Introduction to Quantitative Genetics. Pearson Education; 1996.

5. Lynch M, Walsh B. Genetics and analysis of quantitative traits. Sinauer Associates; 1998.

6. Risch N, Merikangas K. The future of genetic studies of complex human diseases. Science. 1996;273(5281):1516–7.

7. Choi SW, Mak TSH, O’Reilly PF. Tutorial: A guide to performing polygenic risk score analyses. Nature Protocols. 2020;15(9):2759–72.

8. Consortium IS, et al. Common polygenic variation contributes to risk of schizophrenia that overlaps with bipolar disorder. Nature. 2009;460(7256):748.

9. Dudbridge F. Power and predictive accuracy of polygenic risk scores. PLoS Genetics. 2013;9(3):e1003348.

10. Privé F, Vilhjálmsson BJ, Aschard H, Blum MG. Making the most of clumping and thresholding for polygenic scores. The American journal of human genetics. 2019;105(6):1213–21.

11. Mak TSH, Porsch RM, Choi SW, Zhou X, Sham PC. Polygenic scores via penalized regression on summary statistics. Genetic Epidemiology. 2017;41(6):469–80.

12. Ge T, Chen CY, Ni Y, Feng YCA, Smoller JW. Polygenic prediction via Bayesian regression and continuous shrinkage priors. Nature Communications. 2019;10:1776.

13. Lloyd-Jones LR, Zeng J, Sidorenko J, Yengo L, Moser G, Kemper KE, et al. Improved polygenic prediction by Bayesian multiple regression on summary statistics. Nature Communications. 2019;10:5086.

14. Privé F, Arbel J, Vilhjálmsson BJ. LDpred2: better, faster, stronger. Bioinformatics. 2020;36(22-23):5424–31.

15. Zheng Z, Liu S, Sidorenko J, Wang Y, Lin T, Yengo L, et al. Leveraging functional genomic annotations and genome coverage to improve polygenic prediction of complex traits within and between ancestries. Nature Genetics. 2024;56(5):767–77.

16. Mackay TF. The genetic architecture of quantitative traits. Annual review of genetics. 2001;35(1):303–39.

17. Frascaroli E, Cane MA, Landi P, Pea G, Gianfranceschi L, Villa M, et al. Classical genetic and quantitative trait loci analyses of heterosis in a maize hybrid between two elite inbred lines. Genetics. 2007;176(1):625–44.

18. Cui L, Yang B, Xiao S, Gao J, Baud A, Graham D, et al. Dominance is common in mammals and is associated with trans-acting gene expression and alternative splicing. Genome Biology. 2023;24(1):215.

19. Zhu Z, Bakshi A, Vinkhuyzen AAE, Hemani G, Lee SH, Nolte IM, et al. Dominance genetic variation contributes little to the missing heritability for human complex traits. The American Journal of Human Genetics. 2015;96(3):377–85. Available from: https://doi.org/10.1016/j.ajhg.2015.01.001. doi:10.1016/j.ajhg.2015.01.001.

20. Pazokitoroudi A, Chiu AM, Burch KS, Pasaniuc B, Sankararaman S. Quantifying the contribution of dominance deviation effects to complex trait variation in biobank-scale data. The American Journal of Human Genetics. 2021;108(5):799–808. Available from: https://doi.org/10.1016/j.ajhg.2021.03.018. doi:10.1016/j.ajhg.2021.03.018.

21. Guindo-Martínez M, Amela R, Bonàs-Guarch S, Puiggròs M, Salvoro C, Miguel-Escalada I, et al. The impact of non-additive genetic associations on age-related complex diseases. Nature Communications. 2021;12(1):2436. Available from: https://doi.org/10.1038/s41467-021-21952-4. doi:10.1038/s41467-021-21952-4.

22. Palmer DS, Zhou W, Abbott L, Wigdor EM, Baya N, Churchhouse C, et al. Analysis of genetic dominance in the UK Biobank. Science. 2023;379(6639):1341–8.

23. Huang W, Mackay TF. The genetic architecture of quantitative traits cannot be inferred from variance component analysis. PLoS genetics. 2016;12(11):e1006421.

24. Liu Y, et al. Including dominance effects in the prediction model through locus-specific weights on heterozygous genotypes can greatly improve genomic predictive abilities. Heredity. 2022;128:154–66. doi:10.1038/s41437-022-00504-6.

25. Pozarickij A, Williams C, Guggenheim JA. Non-additive (dominance) effects of genetic variants associated with refractive error and myopia. Molecular Genetics and Genomics. 2020;295(4):843–53. Available from: https://doi.org/10.1007/s00438-020-01666-w. doi:10.1007/s00438-020-01666-w.

26. Lu T, Forgetta V, Richards JB, Greenwood CMT. Genetic determinants of polygenic prediction accuracy within a population. Genetics. 2022;222(4):iyac158. Available from: https://doi.org/10.1093/genetics/iyac158. doi:10.1093/genetics/iyac158.

27. Ohta R, Tanigawa Y, Suzuki Y, Kellis M, Morishita S. A polygenic score method boosted by non-additive models. Nature Communications. 2024;15(1):4433. Available from: https://doi.org/10.1038/s41467-024-48654-x. doi:10.1038/s41467-024-48654-x.

28. Hu J, Chen R, Salvatore M, Wu O, Ozdemir OB, Lu Y, et al. A pre-train and fine-tune framework for adaptive boosting of pre-trained polygenic risk scores. Nature Communications. 2026. doi:10.1038/s41467-026-77128-5.

29. Bycroft C, Freeman C, Petkova D, Band G, Elliott LT, Sharp K, et al. The UK Biobank resource with deep phenotyping and genomic data. Nature. 2018;562(7726):203–9.

30. Satterthwaite FE. An approximate distribution of estimates of variance components. Biometrics bulletin. 1946;2(6):110–4.

31. Chang CC, Chow CC, Tellier LCAM, Vattikuti S, Purcell SM, Lee JJ. Second-generation PLINK: rising to the challenge of larger and richer datasets. GigaScience. 2015;4:7.

32. Zou Y, Carbonetto P, Xie D, Wang G, Stephens M. Fast and flexible joint fine-mapping of multiple traits via the Sum of Single Effects model. Nature Genetics. 2026.

33. Yengo L, Sidorenko J, Kemper KE, Zheng Z, Wood AR, Weedon MN, et al. Meta-analysis of genome-wide association studies for height and body mass index in 700000 individuals of European ancestry. Human molecular genetics. 2018;27(20):3641–9.

34. Yengo L, Vedantam S, Marouli E, Sidorenko J, Bartell E, Sakaue S, et al. A saturated map of common genetic variants associated with human height. Nature. 2022;610(7933):704–12.

35. Smit RA, Wade KH, Hui Q, Arias JD, Yin X, Christiansen MR, et al. Polygenic prediction of body mass index and obesity through the life course and across ancestries. Nature medicine. 2025;31(9):3151–68.

36. Hof JP, Speed D. LDAK-KVIK performs fast and powerful mixed-model association analysis of quantitative and binary phenotypes. Nature Genetics. 2025;57(9):2116–23. doi:10.1038/s41588-025-02286-z.

37. Hill WG, Goddard ME, Visscher PM. Data and theory point to mainly additive genetic variance for complex traits. PLoS genetics. 2008;4(2):e1000008.

38. Kunkel D, Sørensen P, Shankar V, Morgante F. Improving polygenic prediction from summary data by learning patterns of effect sharing across multiple phenotypes. PLoS Genetics. 2025;21(1):e1011519.

39. Menzel S, Jiang J, Silver N, Gallagher J, Cunningham J, Surdulescu G, et al. The HBS1L–MYB intergenic region on chromosome 6q23.3 influences erythrocyte, platelet, and monocyte counts in humans. Blood. 2007;110(10):3624–6. doi:10.1182/blood-2007-05-093419.

40. Kullo IJ, Ding K, Jouni H, Smith CY, Chute CG. A genome-wide association study of red blood cell traits using the electronic medical record. PLoS ONE. 2010;5(9):e13011. doi:10.1371/journal.pone.0013011.

41. Stadhouders R, Aktuna S, Thongjuea S, et al. HBS1L-MYB intergenic variants modulate fetal hemoglobin via long-range MYB enhancers. The Journal of Clinical Investigation. 2014;124(4):1699–710. doi:10.1172/JCI71520.

42. Sigurdsson AI, Gräf JF, Yang Z, et al. Complex genetic effects linked to plasma protein abundance in the UK Biobank. Nature Communications. 2026;17:533. doi:10.1038/s41467-025-67235-0.

