## Supplemental Text and Figures for "Accounting for gene action improves the accuracy of polygenic scores for some complex traits"

### Accounting for gene action improves the accuracy of polygenic scores for some complex traits—Supplementary Text and Figures

Yuxuan Sun[1,2], Fabio Morgante[1,3,†,*], Trudy F. C. Mackay[1,3,†,*]

***1*** *Institute for Human Genetics, Clemson University, Greenwood, South Carolina, United States of America*
***2*** *School of Computing, Clemson University, Clemson, South Carolina, United States of America*
***3*** *Department of Genetics and Biochemistry, Clemson University, Clemson, South Carolina, United States of America*
† *These authors jointly supervised the work.*

#### Supplementary Text – S1 Text

##### *Derivation of the test statistics*

Additive ($a$) and dominance ($d$) effects were estimated from genotype-specific phenotype means. Let $n_{\mathrm{A_1A_1}}$, $n_{\mathrm{A_1A_2}}$, and $n_{\mathrm{A_2A_2}}$ denote the sample sizes, $\bar{y}_{\mathrm{A_1A_1}}$, $\bar{y}_{\mathrm{A_1A_2}}$, and $\bar{y}_{\mathrm{A_2A_2}}$ the phenotypic means, and $s^2_{\mathrm{A_1A_1}}$, $s^2_{\mathrm{A_1A_2}}$, and $s^2_{\mathrm{A_2A_2}}$ the phenotypic variances for genotypes $\mathrm{A_1A_1}$, $\mathrm{A_1A_2}$, and $\mathrm{A_2A_2}$, respectively. The additive effect estimator was defined as:

$$\hat{a} = \frac{\bar{y}_{\mathrm{A_1A_1}} - \bar{y}_{\mathrm{A_2A_2}}}{2}. \tag{1}$$

Assuming independence of the phenotypic means across genotypes, the variance of $\hat{a}$ is:

$$\begin{aligned}
\mathrm{Var}(\hat{a}) &= \mathrm{Var}\left(\frac{\bar{y}_{\mathrm{A_1A_1}} - \bar{y}_{\mathrm{A_2A_2}}}{2}\right) \\
&= \frac{1}{4}\mathrm{Var}\left(\bar{y}_{\mathrm{A_1A_1}} - \bar{y}_{\mathrm{A_2A_2}}\right) \\
&= \frac{1}{4}\left[\mathrm{Var}(\bar{y}_{\mathrm{A_1A_1}}) + \mathrm{Var}(\bar{y}_{\mathrm{A_2A_2}})\right] \\
&= \frac{1}{4}\left(\frac{s^2_{\mathrm{A_1A_1}}}{n_{\mathrm{A_1A_1}}} + \frac{s^2_{\mathrm{A_2A_2}}}{n_{\mathrm{A_2A_2}}}\right). \qquad (2)
\end{aligned}$$

The corresponding standard error is:

$$\mathrm{SE}(\hat{a}) = \sqrt{\mathrm{Var(\hat{a})}} = \sqrt{\frac{1}{4}\left(\frac{s^2_{\mathrm{A_1A_1}}}{n_{\mathrm{A_1A_1}}} + \frac{s^2_{\mathrm{A_2A_2}}}{n_{\mathrm{A_2A_2}}}\right)}. \tag{3}$$

The dominance effect estimator was defined as:

$$\hat{d} = \bar{y}_{\mathrm{A_1A_2}} - \frac{\bar{y}_{\mathrm{A_1A_1}} + \bar{y}_{\mathrm{A_2A_2}}}{2}. \tag{4}$$

Assuming independence of the phenotypic means across genotypes, he variance of $\hat{d}$ is

$$\begin{aligned}\mathrm{Var}(\hat{d}) &= \mathrm{Var}\left(\bar{y}_{\mathrm{A_1A_2}} - \frac{\bar{y}_{\mathrm{A_1A_1}} + \bar{y}_{\mathrm{A_2A_2}}}{2}\right) \\ &= \mathrm{Var}(\bar{y}_{\mathrm{A_1A_2}}) + \frac{1}{4}\mathrm{Var}\left(\bar{y}_{\mathrm{A_1A_1}} + \bar{y}_{\mathrm{A_2A_2}}\right) \\ &= \mathrm{Var}(\bar{y}_{\mathrm{A_1A_2}}) + \frac{1}{4}\left[\mathrm{Var}(\bar{y}_{\mathrm{A_1A_1}}) + \mathrm{Var}(\bar{y}_{\mathrm{A_2A_2}})\right] \\ &= \frac{s^2_{\mathrm{A_1A_2}}}{n_{\mathrm{A_1A_2}}} + \frac{1}{4}\left(\frac{s^2_{\mathrm{A_1A_1}}}{n_{\mathrm{A_1A_1}}} + \frac{s^2_{\mathrm{A_2A_2}}}{n_{\mathrm{A_2A_2}}}\right). \end{aligned} \tag{5}$$

The corresponding standard error is:

$$\mathrm{SE}(\hat{d}) = \sqrt{\mathrm{Var}(\hat{d})} = \sqrt{\frac{s^2_{\mathrm{A_1A_2}}}{n_{\mathrm{A_1A_2}}} + \frac{1}{4}\left(\frac{s^2_{\mathrm{A_1A_1}}}{n_{\mathrm{A_1A_1}}} + \frac{s^2_{\mathrm{A_2A_2}}}{n_{\mathrm{A_2A_2}}}\right)}. \tag{6}$$

Statistical significance was evaluated using the following $t$-statistics:

$$t_a = \frac{\hat{a}}{\mathrm{SE}(\hat{a})} = \frac{\dfrac{\bar{y}_{\mathrm{A_1A_1}} - \bar{y}_{\mathrm{A_2A_2}}}{2}}{\sqrt{\dfrac{1}{4}\left(\dfrac{s^2_{\mathrm{A_1A_1}}}{n_{\mathrm{A_1A_1}}} + \dfrac{s^2_{\mathrm{A_2A_2}}}{n_{\mathrm{A_2A_2}}}\right)}}, \tag{7}$$

$$t_d = \frac{\hat{d}}{\mathrm{SE}(\hat{d})} = \frac{\bar{y}_{\mathrm{A_1A_2}} - \dfrac{\bar{y}_{\mathrm{A_1A_1}} + \bar{y}_{\mathrm{A_2A_2}}}{2}}{\sqrt{\dfrac{s^2_{\mathrm{A_1A_2}}}{n_{\mathrm{A_1A_2}}} + \dfrac{1}{4}\left(\dfrac{s^2_{\mathrm{A_1A_1}}}{n_{\mathrm{A_1A_1}}} + \dfrac{s^2_{\mathrm{A_2A_2}}}{n_{\mathrm{A_2A_2}}}\right)}}. \tag{8}$$

Degrees of freedom were approximated using the Welch–Satterthwaite formula [1] to account for unequal variances across genotype groups as:

$$df = \frac{\left(\sum_{k=1}^{K} V_k\right)^2}{\sum_{k=1}^{K} \frac{V_k^2}{df_k}}. \tag{9}$$

Here, $V_k$ denotes the $k$th estimated variance component and $df_k$ its corresponding degrees of freedom. Thus, for the additive effect estimator ($\hat{a}$), the two variance components are:

$$V_{\mathrm{A_1A_1}} = \frac{s^2_{\mathrm{A_1A_1}}}{4n_{\mathrm{A_1A_1}}}, \qquad V_{\mathrm{A_2A_2}} = \frac{s^2_{\mathrm{A_2A_2}}}{4n_{\mathrm{A_2A_2}}}$$

with corresponding degrees of freedom $n_{\mathrm{A_1A_1}} - 1$ and $n_{\mathrm{A_2A_2}} - 1$. Substituting these components into the Welch–Satterthwaite equation gives:

$$df_a = \frac{\left(\frac{s^2_{\mathrm{A_1A_1}}}{n_{\mathrm{A_1A_1}}} + \frac{s^2_{\mathrm{A_2A_2}}}{n_{\mathrm{A_2A_2}}}\right)^2}{\frac{\left(s^2_{\mathrm{A_1A_1}}/n_{\mathrm{A_1A_1}}\right)^2}{n_{\mathrm{A_1A_1}}-1} + \frac{\left(s^2_{\mathrm{A_2A_2}}/n_{\mathrm{A_2A_2}}\right)^2}{n_{\mathrm{A_2A_2}}-1}}. \tag{10}$$

For the dominance effect estimator ($\hat{d}$), the three variance components are

$$V_{\mathrm{A_1A_2}} = \frac{s^2_{\mathrm{A_1A_2}}}{n_{\mathrm{A_1A_2}}}, \qquad V_{\mathrm{A_1A_1}} = \frac{s^2_{\mathrm{A_1A_1}}}{4n_{\mathrm{A_1A_1}}}, \qquad V_{\mathrm{A_2A_2}} = \frac{s^2_{\mathrm{A_2A_2}}}{4n_{\mathrm{A_2A_2}}}$$

with corresponding degrees of freedom $n_{\mathrm{A_1A_2}} - 1$, $n_{\mathrm{A_1A_1}} - 1$, and $n_{\mathrm{A_2A_2}} - 1$. Substituting these components into the Welch–Satterthwaite equation gives:

$$df_d = \frac{\left(\frac{s^2_{\mathrm{A_1A_2}}}{n_{\mathrm{A_1A_2}}} + \frac{s^2_{\mathrm{A_1A_1}}}{4n_{\mathrm{A_1A_1}}} + \frac{s^2_{\mathrm{A_2A_2}}}{4n_{\mathrm{A_2A_2}}}\right)^2}{\frac{\left(s^2_{\mathrm{A_1A_2}}/n_{\mathrm{A_1A_2}}\right)^2}{n_{\mathrm{A_1A_2}}-1} + \frac{\left(s^2_{\mathrm{A_1A_1}}/(4n_{\mathrm{A_1A_1}})\right)^2}{n_{\mathrm{A_1A_1}}-1} + \frac{\left(s^2_{\mathrm{A_2A_2}}/(4n_{\mathrm{A_2A_2}})\right)^2}{n_{\mathrm{A_2A_2}}-1}}. \tag{11}$$

To confirm that the test statistics behaved as expected under the null hypothesis, we performed 10,000 independent simulation replicates, each consisting of a single genetic variant and 50,000 individuals, with allele frequencies randomly sampled between 0.05 and 0.95 (*i.e.*, excluding variants with MAF $< 0.05$). Under the null hypothesis of no additive or dominance effects of genotype ($a = d = 0$), we generated phenotypes from normal distribution with a mean of 0, allowing phenotype variances to differ across the three genotype groups by independently sampling genotype-specific variances from a uniform distribution, $U(0.5, 5)$, for each replicate. We then estimated $\hat{a}$ and $\hat{d}$ from genotype-specific phenotype means and computed test statistics and degrees of freedom using equations 1, 4, 7, 8, 10, and 11, respectively. The empirical distributions of the resulting test statistics closely followed the theoretical $t$-distribution quantiles (S1 Figure). This result confirms that the variance estimators and corresponding $t$-statistics are well calibrated and that the $t$-distribution assumption holds under the null hypothesis of $a = 0$ and $d = 0$.

#### Supplementary Figures

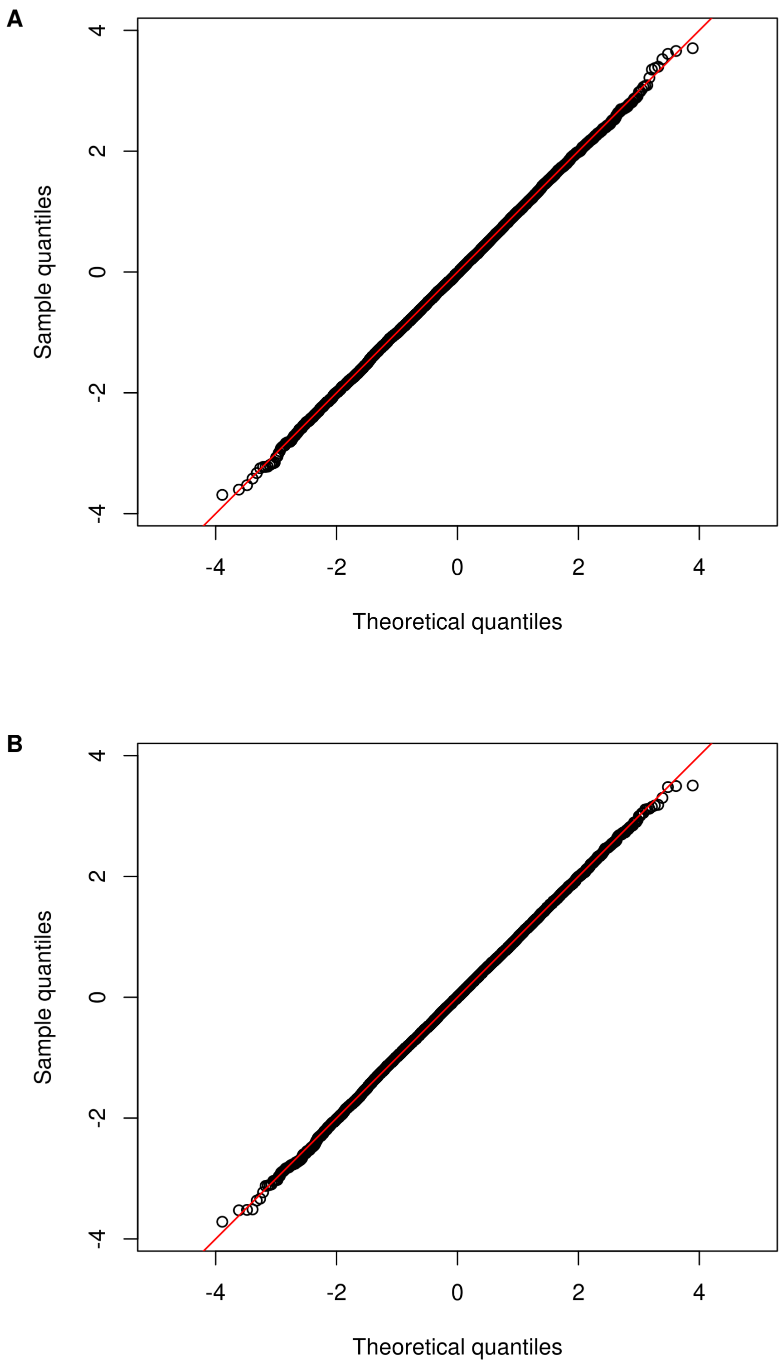


**S1 Fig. Q-Q plots of the test statistics for estimated additive and dominance effects under the null hypothesis.** The empirical distributions of the test statistics are compared with the theoretical $t$-distribution quantiles for (A) the estimated additive effect ($\hat{a}$) and (B) the estimated dominance effect ($\hat{d}$). The test statistics closely follow the theoretical quantiles and align along the diagonal, indicating that the variance estimators and corresponding $t$-statistics are well calibrated under the null hypotheses $a = 0$ and $d = 0$.

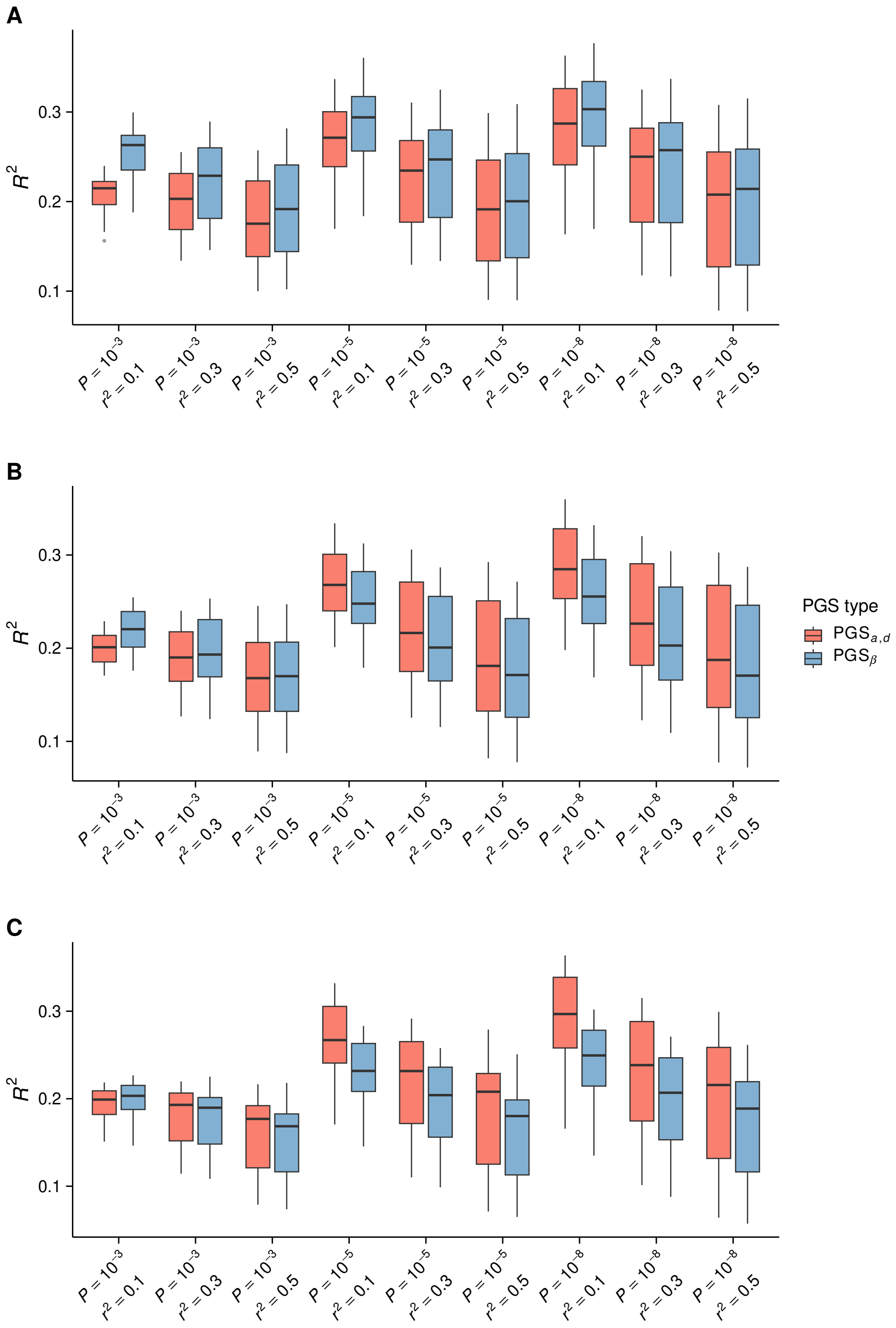


**S2 Fig. Comparison of $R^2$ between PGS$_{a,d}$ and PGS$_\beta$ across parameter combinations and genetic architectures.** Prediction accuracy was evaluated using $R^2$ across combinations of clumping *P*-value thresholds ($P < 10^{-3}$, $10^{-5}$, and $10^{-8}$) and LD $r^2$ thresholds ($r^2 = 0.1$, $0.3$, and $0.5$) under three simulated genetic architectures. (A) Additive architecture. (B) Combined additive and dominant architecture. (C) Dominant architecture. Model performance was summarized across simulation replicates for each parameter combination.

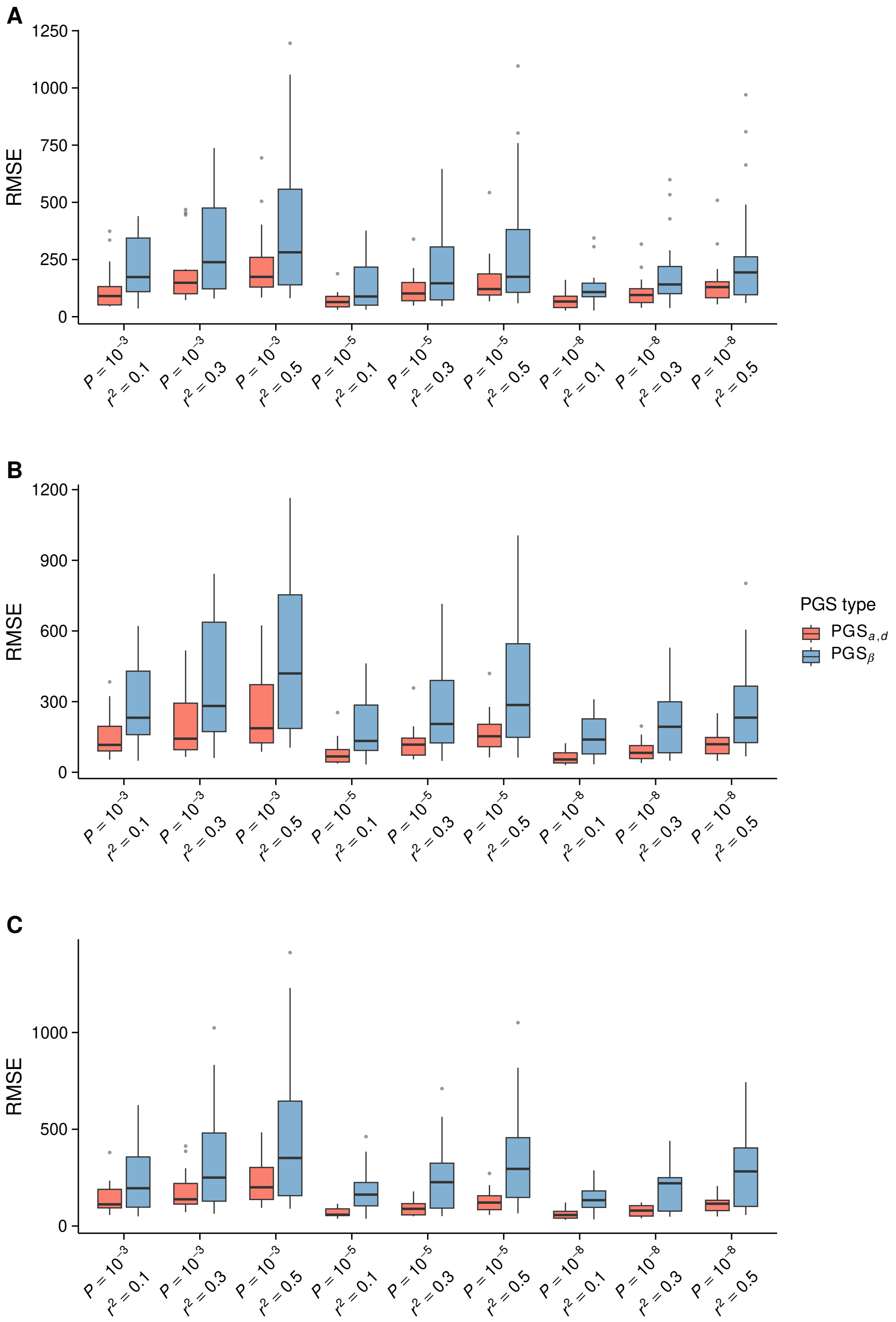


**S3 Fig. Comparison of RMSE between PGS$_{a,d}$ and PGS$_{\beta}$ across parameter combinations and genetic architectures.** Prediction accuracy was evaluated using the root mean squared error (RMSE) across combinations of clumping *P*-value thresholds ($P < 10^{-3}$, $10^{-5}$, and $10^{-8}$) and LD $r^2$ thresholds ($r^2 = 0.1$, $0.3$, and $0.5$) under three simulated genetic architectures. (A) Additive architecture. (B) Combined additive and dominant architecture. (C) Dominant architecture. Model performance was summarized across simulation replicates for each parameter combination.

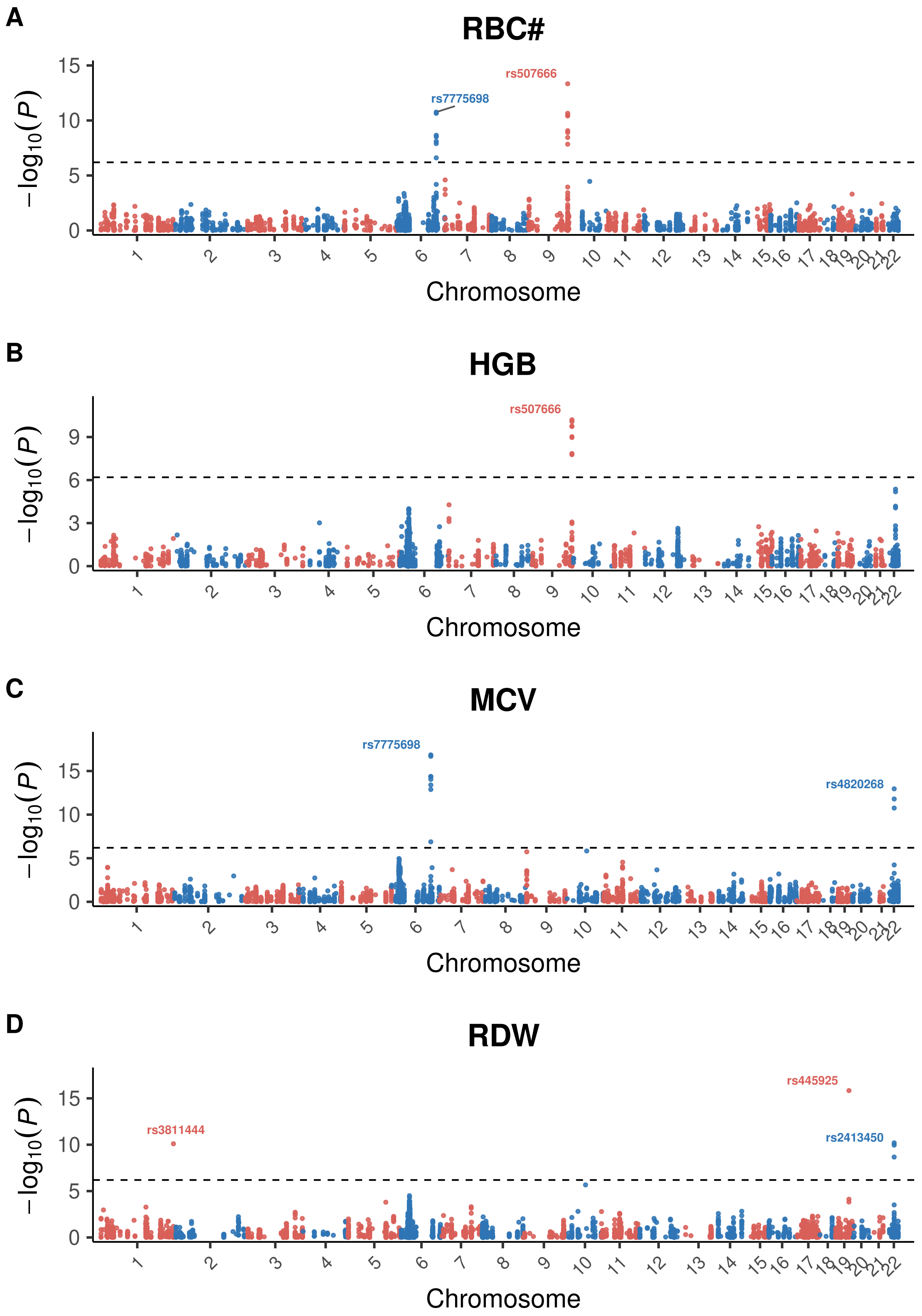
A
RBC#
rs507666
rs7775698
B
HGB
rs507666
C
MCV
rs7775698
rs4820268
D
RDW
rs445925
rs3811444
rs2413450
$-\log_{10}(P)$
Chromosome

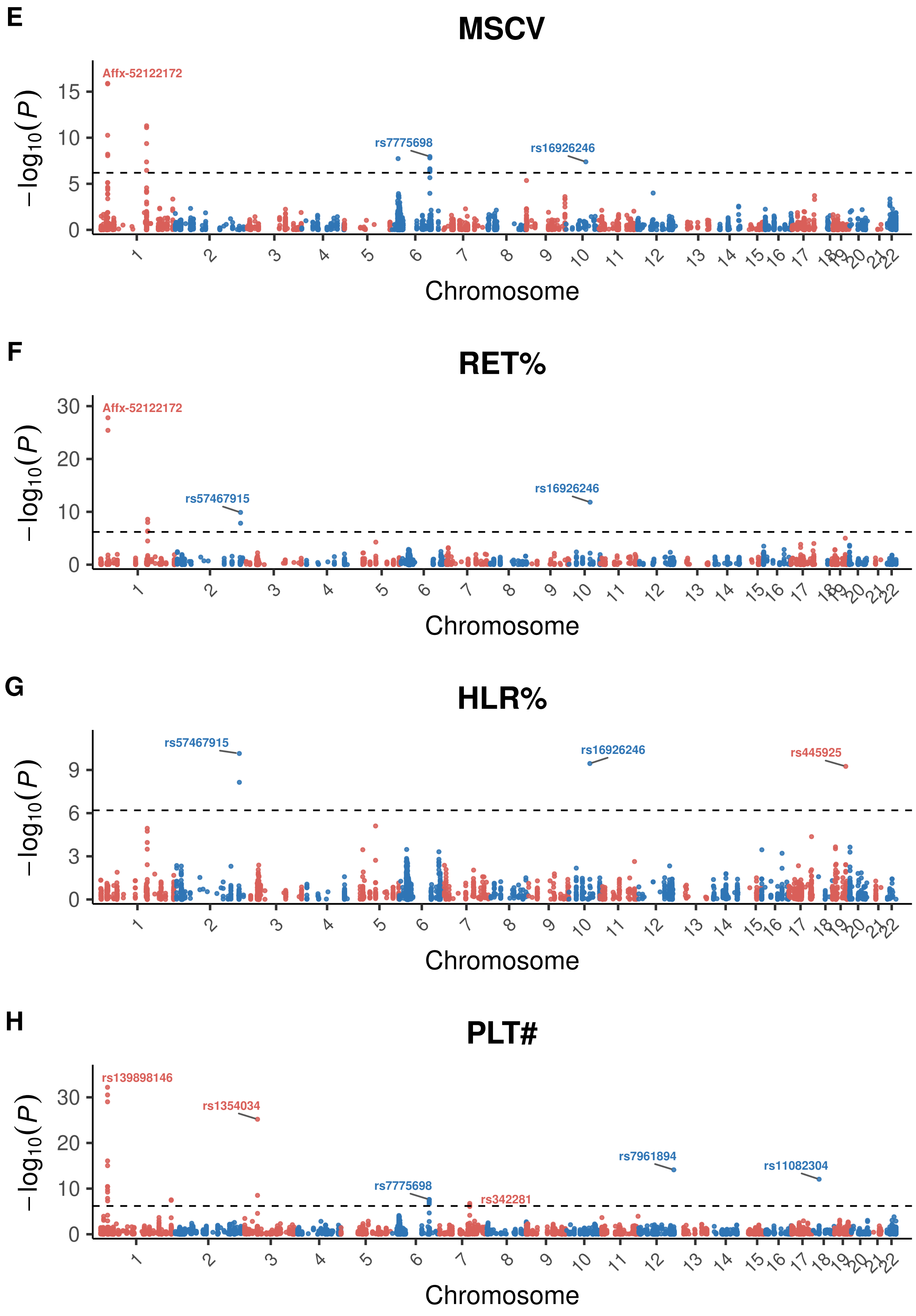
E
MSCV
Affx-52122172
rs7775698
rs16926246
F
RET%
Affx-52122172
rs57467915
rs16926246
G
HLR%
rs57467915
rs16926246
rs445925
H
PLT#
rs139898146
rs1354034
rs7961894
rs11082304
rs7775698
rs342281
−log10(P)
Chromosome

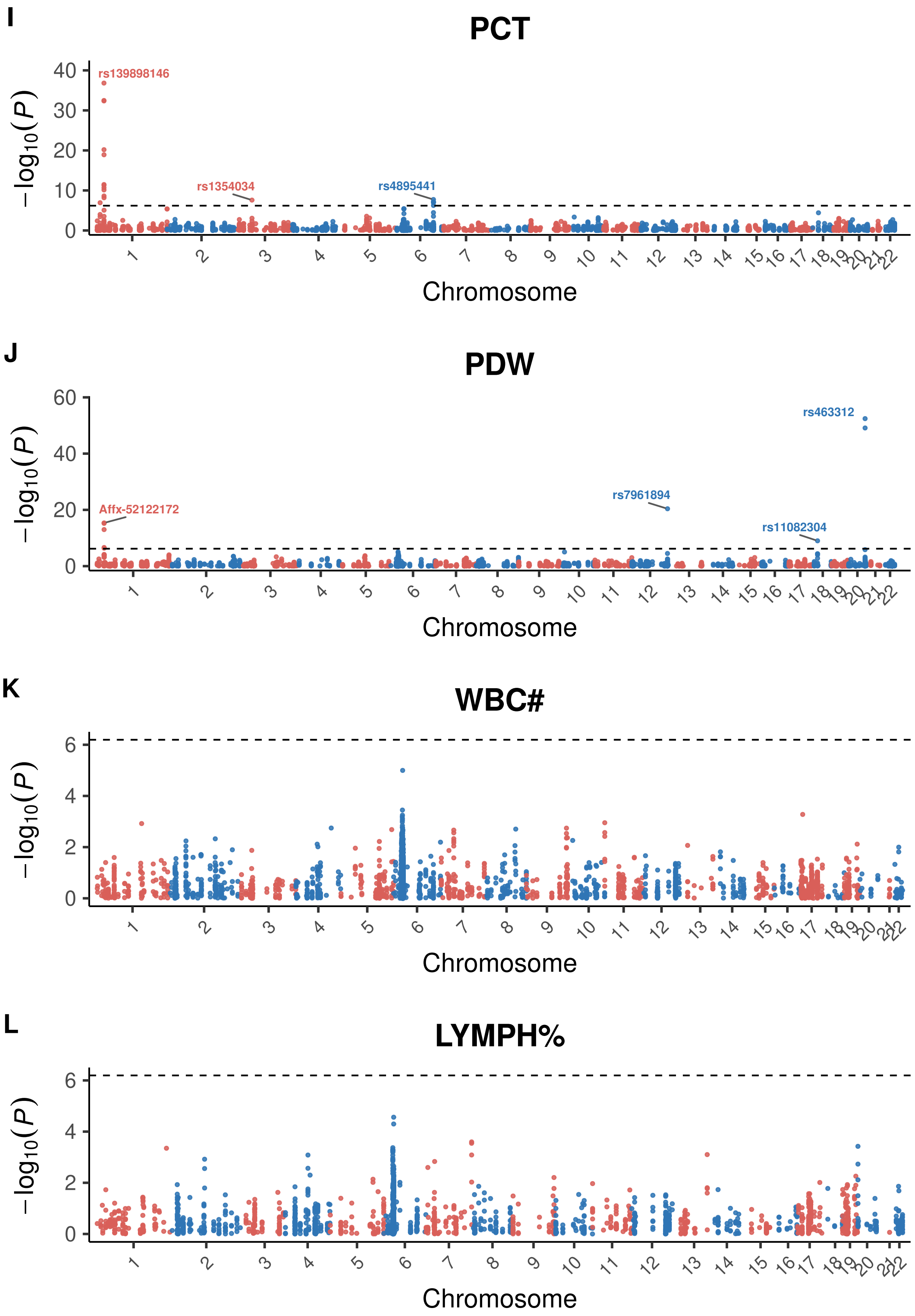
I
PCT
rs139898146
rs1354034
rs4895441
J
PDW
rs463312
rs7961894
Affx-52122172
rs11082304
K
WBC#
L
LYMPH%
$-\log_{10}(P)$
Chromosome

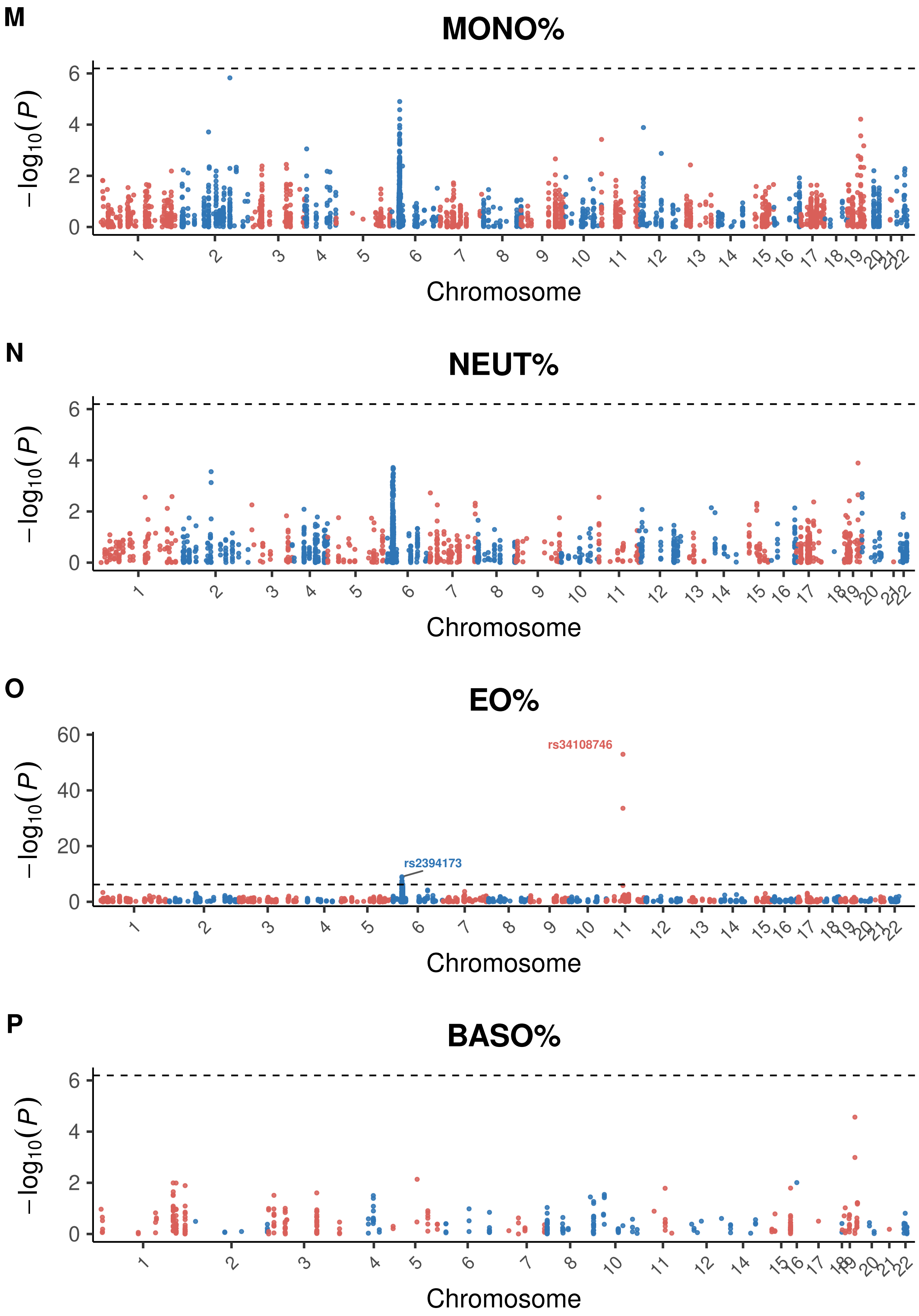
M
MONO%
−log10(P)
Chromosome
N
NEUT%
−log10(P)
Chromosome
O
EO%
rs34108746
rs2394173
−log10(P)
Chromosome
P
BASO%
−log10(P)
Chromosome

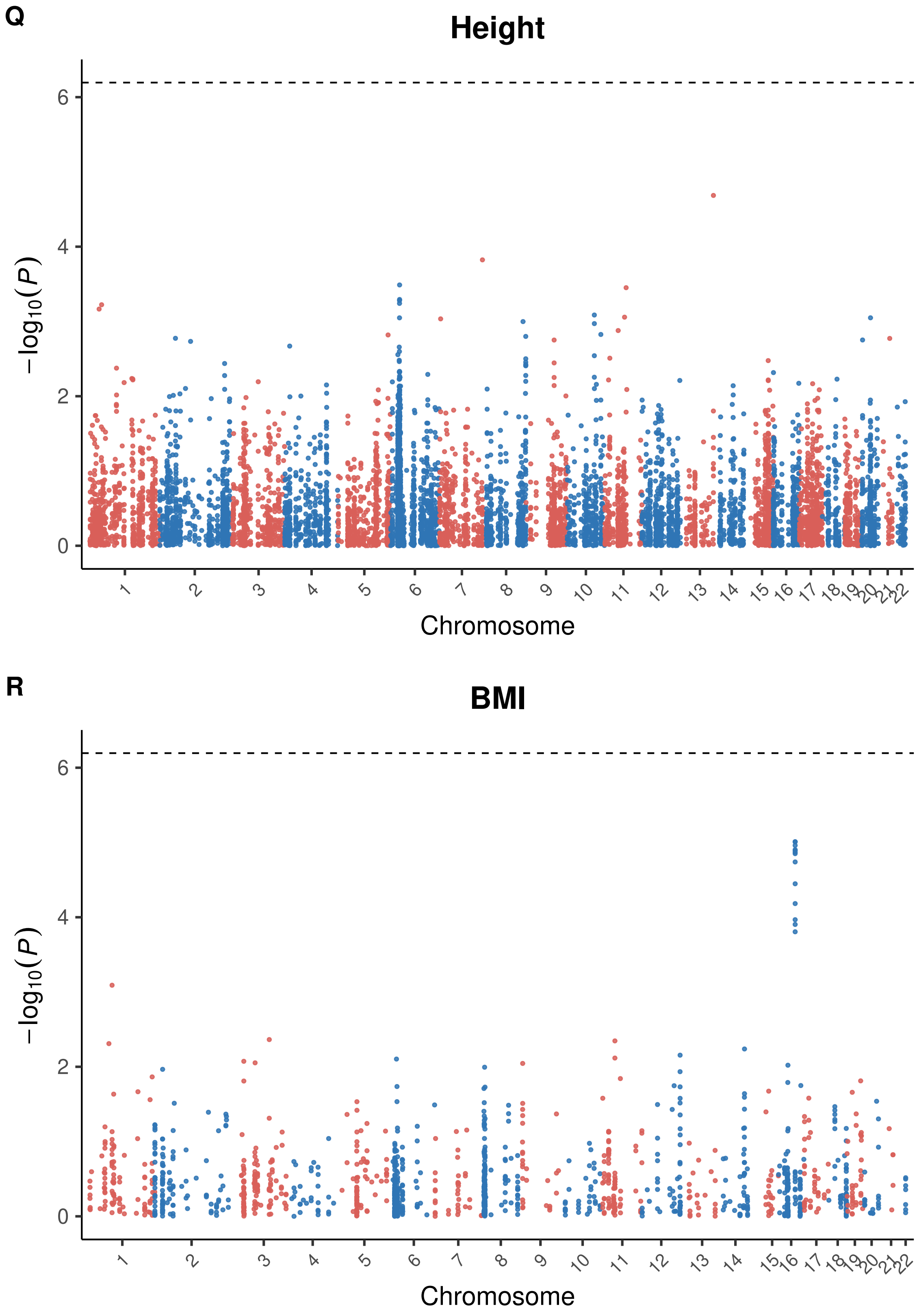


**S4 Fig. Manhattan plots of dominance effects.** Manhattan plots for dominance effects ($\hat{d}$) for variants analyzed across blood cell traits, height, and BMI. Each panel is a different trait and the dashed lines indicate the Bonferroni-corrected significance thresholds.

#### References

[1] F. E. Satterthwaite. An approximate distribution of estimates of variance components. *Biometrics bulletin*, 2(6):110–114, 1946.
